# Cortical representation of sensation elicited by peripheral nerve stimulation in an individual with incomplete spinal cord injury

**DOI:** 10.1101/2025.09.09.25335042

**Authors:** Preethisiri Bhat, William Memberg, Brianna Hutchison, Bryn Spilker, Rohit Bose, Crispin Foli, Aaron Ketting-Olivier, Dawn Taylor, Robert F. Kirsch, Eric Herring, Jennifer A. Sweet, Jonathan Miller, A. Bolu Ajiboye, Emily L. Graczyk

## Abstract

Peripheral nerve stimulation (PNS) is a promising technique for restoring touch to people with neurological injuries. However, its application remains limited because past studies solely focused on people with limb loss and because the optimal paradigm for encoding touch information is still unclear. This study investigated PNS as a modality to restore touch in an individual with sensory-incomplete spinal cord injury (SCI) and quantified intracortical activity in primary somatosensory cortex (S1) resulting from PNS. S1 activity serves as an objective measure to compare and refine PNS paradigms to ultimately improve the effectiveness of PNS as sensory feedback in bidirectional neuroprostheses. We found that PNS delivered to the median and ulnar nerves via chronically-implanted, multi-contact cuff electrodes consistently evoked hand sensations with intensities that reliably scaled with stimulation pulse width (PW). In S1, increasing PNS PW recruited larger cortical populations, increased multi-unit firing rates, and shortened latencies between PNS onset and peak S1 activation. Interestingly, most S1 responses to PNS had strong onset transients, like those previously observed in response to mechanical indentation. Because our PNS paradigm was designed to recruit a population of peripheral afferents synchronously at a fixed frequency, our results suggest that central mechanisms play a role in producing cortical onset transients. This study supports PNS as a viable sensory feedback approach for individuals with incomplete SCI and reveals the representation of electrically-evoked sensory percepts in human cortex for the first time.

## Introduction

Touch is a fundamental sense that shapes how we interact with the world and connect with one another. It provides essential information required for dexterous manipulation and precise grasping of objects^1–3^, and allows us to convey a wide range of emotions to others^4,5^. However, people with spinal cord injury (SCI) can have their sense of touch significantly impaired. Roughly 182,000 individuals living in the United States with an SCI are tetraplegic, resulting in paralysis of the arms and legs and often a reduction or complete loss of touch sensation below the level of injury^6^. Survey studies have shown that regaining arm and hand function is the top priority for improving the quality of life for those living with tetraplegia^7^. However, without touch feedback, hand-object interactions are more difficult to control^1^. Therefore, restoring the sense of touch is a critical aspect of functional restoration for people with tetraplegia.

Intracortical microstimulation (ICMS) has been proposed as a possible technique to provide sensory feedback to people with SCI^8–11^. However, these approaches require complex and invasive brain surgery, and the chronic stabilities of the elicited percepts and the intracortical interface itself are likely limited^12,13^. In contrast, PNS delivered via implanted neural interfaces has demonstrated long-term stability in clinical trials and has restored sensation to people with upper and lower limb amputation for more than a decade^14–22^. PNS can also potentially be translated to the clinic faster than ICMS for people with sensory-incomplete SCIs, which account for nearly half of all SCIs^6^. While the benefits of PNS for motor function after SCI have been well characterized^23,24^, only one study to our knowledge has attempted to use PNS as sensory substitution after SCI^25^.

Prior studies in people with limb loss have demonstrated that manipulating PNS pulse parameters, such as pulse width (PW) and pulse frequency, can modify the perceptual properties of the evoked sensation^19,26–28^. However, traditional PNS is frequently described as ‘paresthetic’ or ‘unnatural’ by participants, and thus may not yield the most natural or informative sensory percepts^29^. In natural touch, rapidly-adapting (RA) fibers respond to the rate of skin indentation but are quiescent during static indentations, while slowly-adapting (SA) fibers remain activated throughout the duration of a constant skin indentation^29,30^. However, unlike these different firing patterns in different afferent fiber types that occur in natural touch, PNS likely recruits a mixed group of both RA and SA fibers, and these recruited fibers likely fire continuously and synchronously for the entire duration that PNS is applied^31^. Studies have shown that neurons in the primary somatosensory cortex (S1) take in inputs from both SA and RA afferent populations during natural touch, resulting in cortical responses that lie on a spectrum between the two firing patterns seen in the periphery^32^. There is little known about how the abnormal recruitment of afferent fibers from PNS affects cortical responses and whether the responses may be able to explain aspects of the stimulation-elicited sensation. One prior study in non-human primates evaluated several stimulation paradigms through cortical recording during PNS^33^, but there have not been any clinical studies yet.

In this study, we take the first step in determining whether PNS could be a viable modality to restore missing sensations in individuals with sensory-incomplete SCI. In addition, we explore cortical measures to investigate what the effect of unnatural recruitment of peripheral fiber types from PNS is in S1. We collected data from a participant with cervical AIS-B SCI enrolled in the Reconnecting the Hand and Arm to the Brain (ReHAB) clinical trial, who was implanted with microelectrode arrays in several regions of the cortical grasp network, including S1, as well as multi-channel nerve cuff electrodes around the contralateral upper extremity nerves, such as the median and ulnar nerves^11^. This system enabled us to record high-resolution intracortical spiking activity in S1 while delivering sensory PNS. We sought to answer three questions. First, given that below-injury PNS has not previously been evaluated as a form of sensory feedback for people with sensory-incomplete SCI, we wished to confirm that below-injury PNS can produce reliable, scalable sensation in this individual. Second, we examined the cortical response to traditional sensory PNS to understand how it compares to the expected peripheral response based on the field’s understanding of peripheral activation patterns. Third, given that this study was conducted with a human participant, we have access to both perceptual and neural responses and thus aimed to understand how cortical metrics correlate to aspects of the perceptual experience. We focused our analyses on the effect of stimulation PW on both the perceptual and cortical response because prior studies have shown that increasing the PNS PW leads to reliable increases in sensation intensity^19^. In the context of neuroprostheses, sensation intensity is critical for conveying how much pressure is being applied on objects by a prosthesis or robotic hand^16,34^.

Answering these questions is the first step in determining whether PNS could provide benefits when integrated into a bi-directional neuroprosthesis for individuals with sensory-incomplete SCI. Additionally, this study will expand our understanding of how sensory signals are transmitted along the sensory neuraxis and represented cortically.

## Results

We characterized the perceptual and cortical response to PNS in a male participant with AIS-B C3/C4 SCI sustained approximately five years prior to enrolling in the ReHAB clinical trial. In this study, PNS was applied to the median and ulnar nerves via the 16-channel Composite Flat Interface Nerve Electrodes (C-FINEs) implanted around these nerves (Figure 1A). PW levels were selected for each block as a percentage of the participant’s sensory dynamic range, which was defined as the range from detection threshold (0% of the dynamic range) to maximum comfortable sensation (100% of the dynamic range) (Figure 1B, C). In each session, PW was pseudorandomly varied over both subthreshold and suprathreshold ranges.

**Figure 1.**
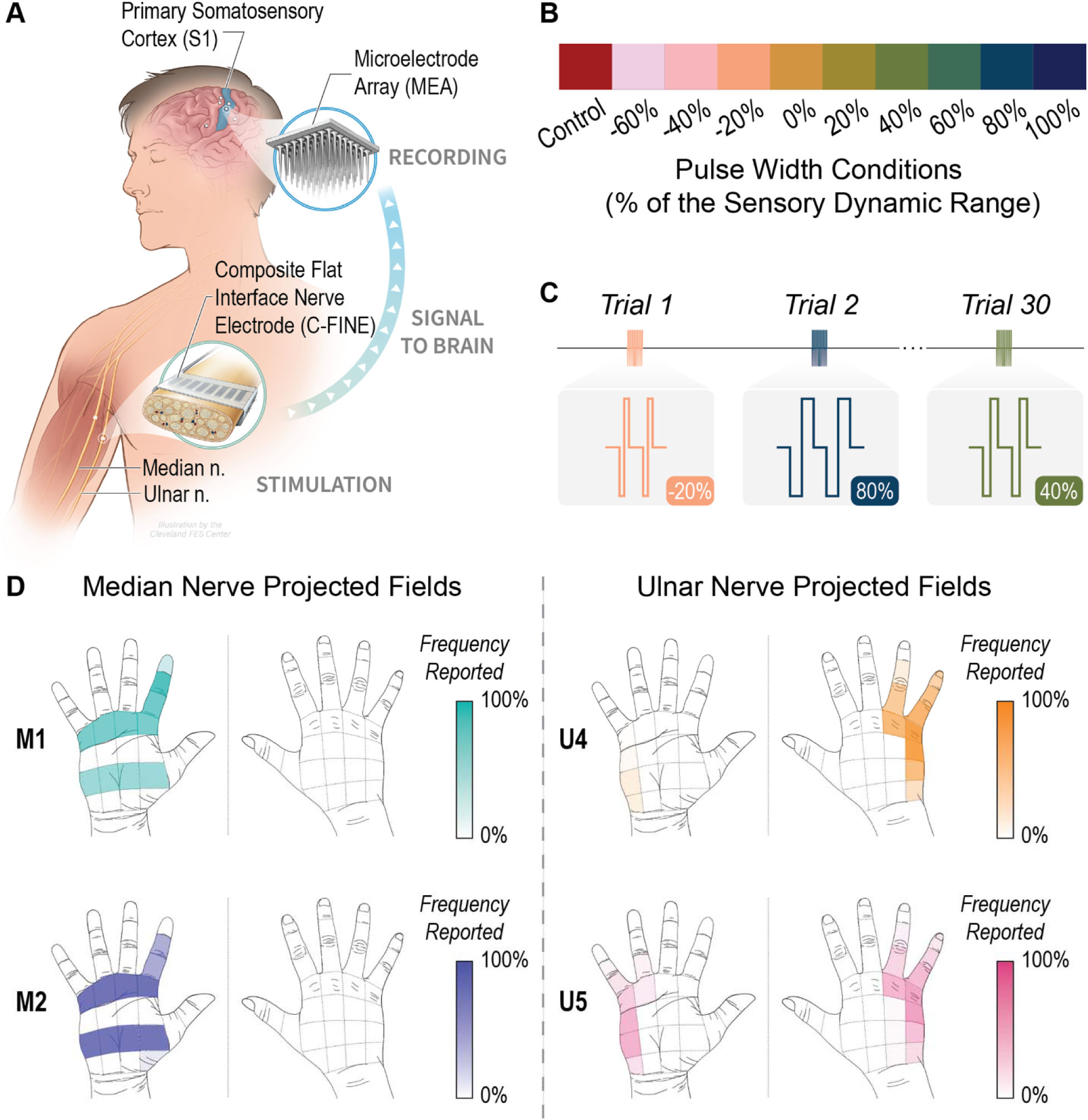
Peripheral nerve stimulation evoked touch sensation on the hand. A) Peripheral nerve stimulation was delivered via two 16-channel Composite Flat Interface Nerve Electrodes (C-FINEs), which were placed around the median and ulnar nerves. Intracortical recordings were made via two 64-channel microelectrode arrays implanted in S1. B) Experimental conditions spanned subthreshold and suprathreshold pulse width values across the sensory dynamic range (-60% to 100%, where 0% is detection threshold and 100% is the maximum comfortable level). The control condition was no stimulation. C) Trial sequence for a given block. Pulse width conditions were pseudorandomly delivered to the participant, and each block consisted of 30 trials. D) Projected fields from PNS delivered to select contacts on the median nerve (marked as M1 and M2) and ulnar nerve (marked as U4 and U5). Opacity of each hand region indicates the percentage of trials in which it was reported across all sessions for the given C-FINE contact.

### PNS evoked sensory percepts on the hand

Since the study participant retained some intact sensation below his injury level (Supplemental Table 1), we first sought to confirm that the participant could perceive PNS delivered to the below-injury nerves. This step was needed to demonstrate that neural activity in the periphery generated by PNS could be relayed to the brain.

As anticipated, delivering PNS to the median and ulnar nerves reliably produced sensory percepts on the hand (Supplemental Figure 1). We selected two contacts per nerve for the remainder of this study. Stimulating the median nerve contacts elicited sensation on the ventral side of the index finger and palm (Figure 1D left, Supplemental Figure 2). In contrast, stimulating the ulnar nerve contacts produced sensation on both the dorsal and ventral sides of the ring and pinky finger and the hypothenar eminence (Figure 1D right, Supplemental Figure 2). The sensation locations elicited for each contact remained consistent throughout the entire duration they were tested (Supplemental Figure 2, Supplemental Figure 3). The sensory percepts generated by PNS were described as ‘buzzing’, ‘tingling’, and ‘electrical’, and this quality remained consistent across all contacts and sessions.

### Perceived intensity increased as stimulation pulse width increased

We next sought to confirm that perceived intensity would scale with PNS pulse width, as has previously been demonstrated in studies of people with limb loss^19^. We found that perceived intensity monotonically increased with PW for the suprathreshold conditions (i.e., 0-100% of the sensory dynamic range) (linear regression t-test, p<0.0001). Consistent with prior PNS studies^19^, a sigmoidal curve provided a good fit to the perceived intensity ratings across PW (average R^2^ of 0.91 +/- 0.06 across sessions) (Figure 2A, B). For each contact tested, the slope of the sigmoidal fits was approximately the same across sessions (Figure 2B). When the intensity ratings across sessions were aggregated together, the means of the normalized perceived intensity ratings per PW condition were significantly different across all the suprathreshold conditions (ANOVA p<0.0001, post-hoc comparisons with Tukey-Kramer test, p<0.0001) (Figure 2C). Subthreshold conditions usually had no sensory percept, and hence, a perceived intensity rating of zero. The distribution of intensity ratings was not significantly influenced by the specific contact or nerve being stimulated, or by the day the sessions were conducted (ANOVA, p>0.05).

**Figure 2.**
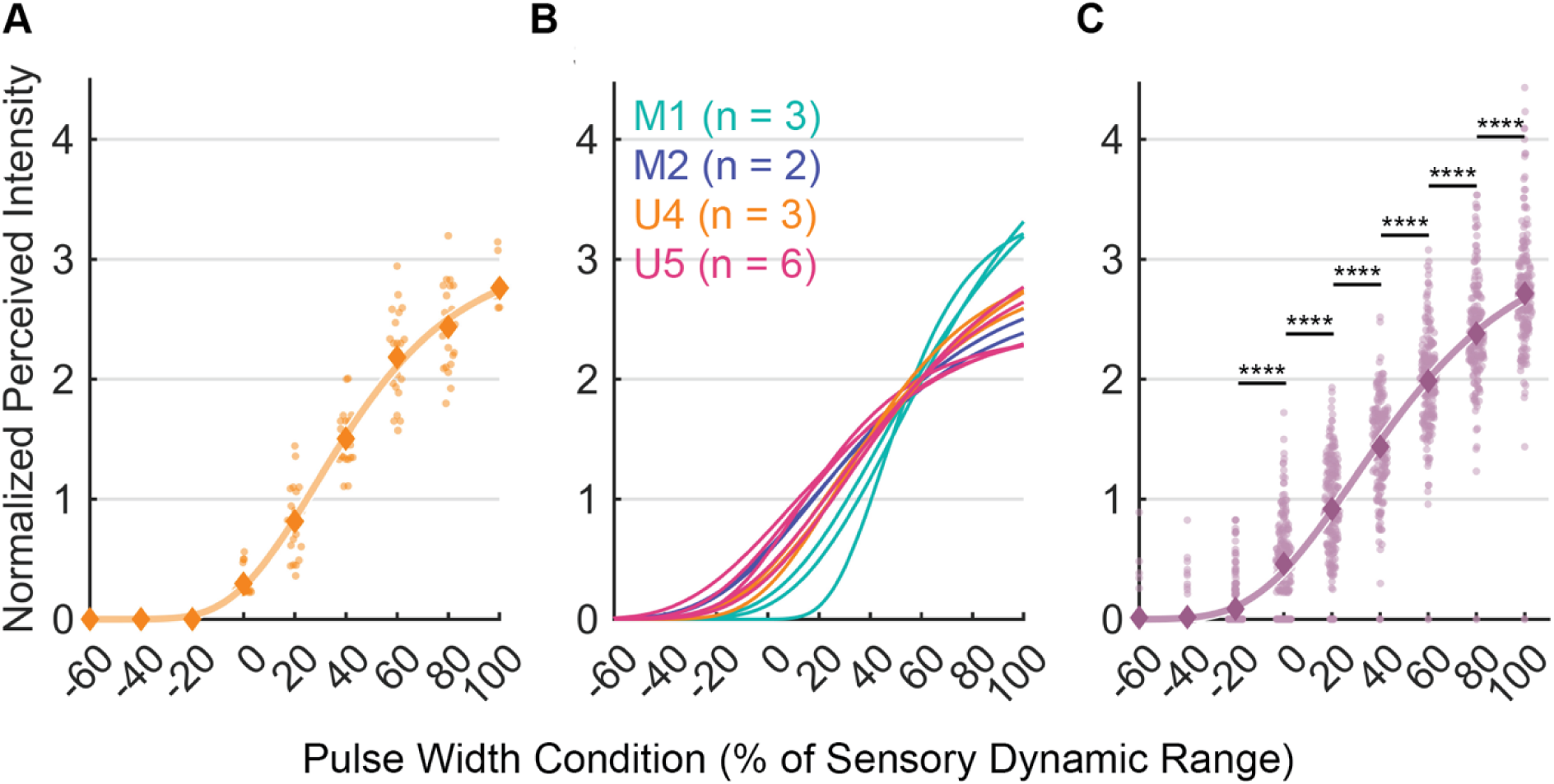
Perceived sensation intensity from below-injury PNS in a participant with sensory-incomplete tetraplegia. A) Example dataset demonstrating how perceived intensity increased with PW condition (% of dynamic range) for contact U4. A sigmoidal curve was fit to the normalized perceived intensity ratings across PW conditions spanning the sensory dynamic range (n = 22 trials per condition). Diamonds indicate the mean intensity for a given PW condition. B) The positive sigmoidal relationship between stimulation PW and perceived intensity was consistent across stimulation contacts and sessions. Each session’s sigmoidal fit is given as a separate line (n = 14 sessions). Colors denote different PNS contacts used for stimulation. C) Normalized perceived intensity ratings aggregated across all sessions (n = 14 sessions, 211-218 trials per PW condition) with sigmoidal fit. Diamonds indicate the mean intensity for each PW condition. **** denote significant differences at p < 0.0001.

### The cortical response to PNS is not a simple aggregation of peripheral neural responses

Given that the participant perceived below-injury PNS, we next sought to determine the cortical representation of PNS in S1. Towards this end, intracortical activity was recorded from two 64-channel Utah microelectrode arrays in area 1 of S1 during all trials^35^ (Figure 1A). Specifically, the arrays were placed in regions of area 1 with projected fields spanning the index, middle, and ring fingers that were found during the awake brain mapping^11^. The cortical response to each PNS stimulus was broken down into several epochs - the onset transient, the sustained response during the stimulus, and the offset transient. Electrodes in S1 were labeled as ‘responsive’ if the normalized firing rate for one or more epochs differed significantly from baseline for at least two suprathreshold conditions. To classify the firing patterns on these electrodes across conditions, we generated an ‘activation profile’ consisting of a timeseries of normalized binned firing rates for each responsive electrode. Only stimulation conditions where the electrode displayed firing significantly different from baseline were included in the subsequent analysis.

The current understanding of PNS activation in the periphery is that PNS synchronously activates a mixed population of afferent fibers (i.e., SAI, RAI, and Pacinian corpuscles)^16,22,31^, where each PNS pulse will induce an action potential in the entire recruited afferent population. Thus, in response to PNS, the recruited afferents will fire at a fixed rate that matches the stimulation frequency throughout the entire period of stimulation (Figure 3A, B) and all recruited peripheral afferents will fire in the same pattern. We hypothesized that the cortical activation pattern from PNS would reflect an aggregation of peripheral afferent firing patterns. Therefore, we expected that activated cortical neurons would display consistently elevated firing throughout the onset and sustained epochs of the PNS stimuli.

**Figure 3.**
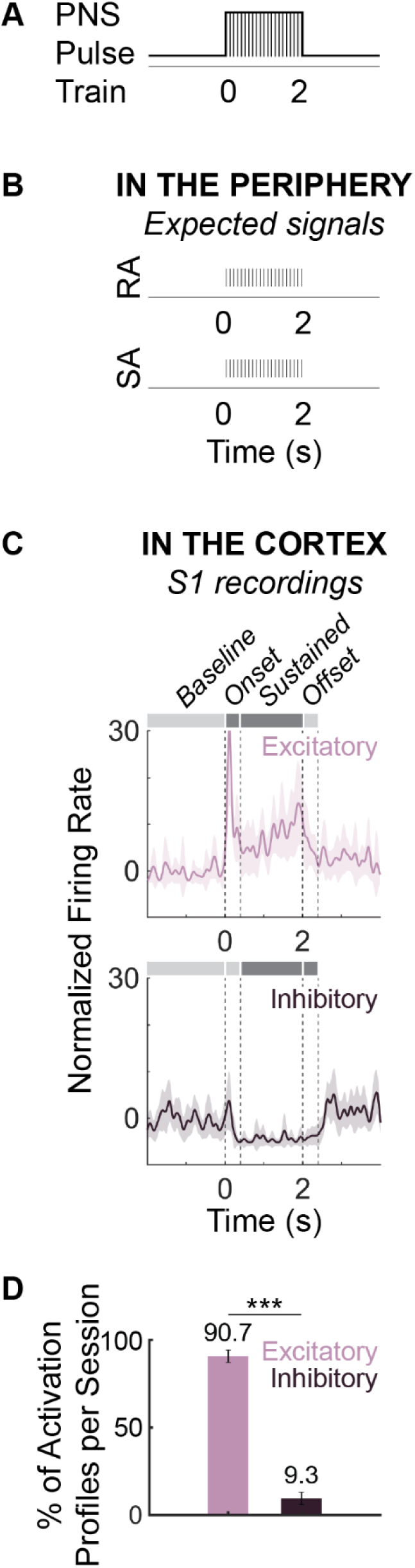
Cortical response to PNS. A) Illustration of the envelope of a two-second PNS pulse train composed of charge-balanced, rectangular pulses with fixed amplitude, width, and frequency. B) Illustration of the expected firing response in the peripheral nerves for a rapidly adapting (RA) and slowly adapting (SA) afferent fiber in response to the two-second PNS pulse train shown in A. Each line represents an action potential. C) Example cortical activation profiles recorded from individual electrodes in the area 1 microelectrode arrays, depicted as peri-stimulus time histograms (PSTHs). Activation profiles were time-binned into three epochs: onset, sustained, and offset. The top PSTH is an example of an excitatory activation profile because the firing rate was significantly higher in the onset and sustained epochs compared to baseline (epochs significantly different from baseline indicated by dark grey). The bottom PSTH is an example of an inhibitory activation profile because the firing rate was significantly lower in the sustained and offset epochs compared to baseline. D) Average percentage of excitatory v. inhibitory activation profiles pooled across responsive PW conditions and electrodes per session (n = 10 sessions, 4-150 activation profiles per session, 573 activation profiles total). Error bars denote the standard error of the mean for the percentages across sessions. ‘***’ denote a significant difference of p < 0.001.

Instead, our data showed that the activation profiles of the responsive cortical electrodes had a strong onset transient, which is qualitatively similar to the cortical response to natural touch in prior NHP studies^35^ (Fig 3C top). Among the responsive electrodes, both excitatory and inhibitory activation profiles were observed in S1 in response to PNS (Figure 3C), but on average, the fraction of excitatory activation profiles per session exceeded the inhibitory activation profiles by almost a factor of 10 (Figure 3D). Therefore, further analysis focused on excitatory activation profiles only.

Out of the excitatory activation profile pool across sessions, on average 91.4% of the activation profiles were responsive during the onset epoch (Figure 4A, B), and 77.7% of all excitatory activation profiles displayed responsivity solely during onset (Figure 4A blue bar, Figure 4B blue profile). The remaining 13.7% of excitatory activation profiles active during onset were also active in one or more additional epochs (Figure 4A, B, green, purple, and brown conditions). The most prevalent types of these activation profiles were those with elevated firing during both the beginning and end of stimulation (i.e., displaying both an onset and offset response, 5.6% of profiles) (Figure 4B, purple) and those active for the entire stimulation duration (i.e., displaying a response during onset and sustained epochs, 4.5% of profiles) (Figure 4B, green). Only 8.6% of all activation profiles were active exclusively in the sustained and/or offset epochs (Figure 4A, yellow, red, and orange bars) and of these, 5.2% of all activation profiles were responsive during only stimulation offset (Figure 4B, red).

**Figure 4.**
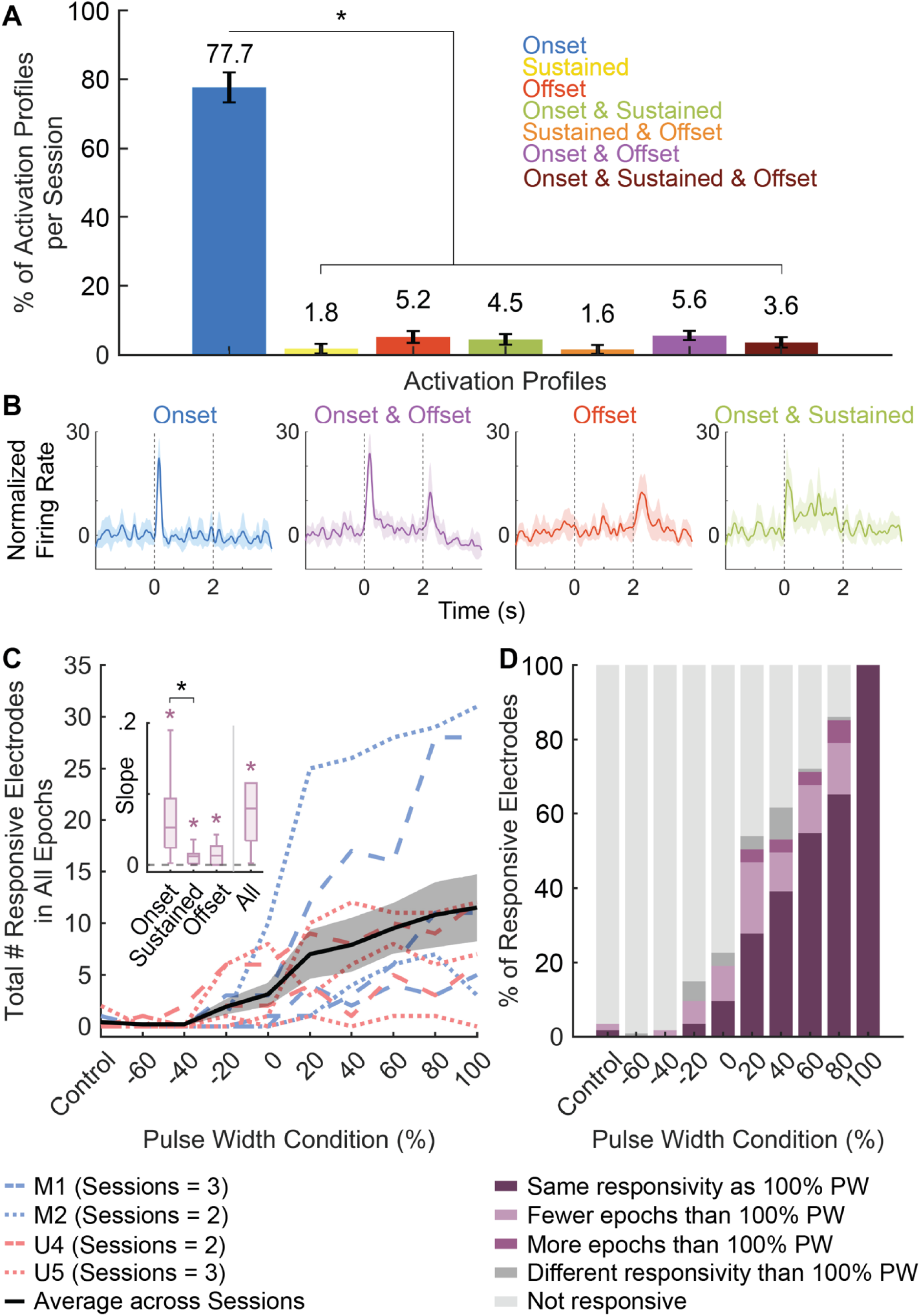
Characterization of excitatory cortical activity across cortical electrodes. A) Average percentages of different excitatory activation profiles pooled across responsive PW conditions and electrodes per session (n = 10 sessions, 525 excitatory activation profiles total). Activation profiles are categorized by the epochs (onset, sustained, and/or offset) that had significantly elevated neural activity compared to baseline. Error bars denote the standard error of the mean of the percentages across sessions. Black ‘*’ denotes significant differences of percentages between activation profile patterns at p < 0.05. B) Example excitatory activation profiles for the four most frequently occurring profiles across sessions: onset only (blue), onset and offset (purple), offset only (red), and offset and sustained (green). Activation profiles are shown as peri-stimulus time histograms (PSTHs). C) The total number of electrodes responsive to PNS in any epoch increased with increasing PW for both median (blue) and ulnar (coral) stimulation contacts. Each patterned line represents a different session, and the pattern of the line represents which contact was being stimulated in the session. The black line is the average number of electrodes responsive in any epoch across all sessions and the shaded region is the standard error of the mean across sessions (n = 10 sessions). Inset: We determined the slope of the relationships between PW condition and the number of responsive electrodes for individual epochs (onset, sustained, and offset). We also included the slope of the relationship between PW condition and the total number of responsive electrodes in any epoch (shown in the main panel). Each boxplot represents the distributions of slopes for the given epoch across all sessions (n = 10 sessions). Purple ‘*’ denote slopes that are significantly different from zero (i.e., no slope) at p < 0.01. The black ‘*’ denotes significant differences of slopes across epochs at p < 0.03. D) Pattern of behavior of the responsive electrodes across PW conditions. The set of electrodes responsive in any epoch at the 100% PW condition were aggregated across sessions (n = 103 electrodes total). The activation profile of each electrode was then compared for each PW condition to its profile at 100% PW within sessions. For each PW condition, the electrode could have the same responsivity in the given PW condition as in the 100% PW condition (dark purple), be responsive in additional epochs at 100% compared to the given PW condition (light purple), be responsive in fewer epochs at 100% compared to the given PW condition (medium purple), be responsive in entirely different epochs from those in 100% PW (dark grey), or be unresponsive (i.e., no activity greater than baseline) at the given PW condition (light grey). Note that the light grey condition is a byproduct of the increase in number of responsive channels with PW shown in panel C.

Interestingly, the strong onset transient observed in the vast majority of activation profiles is inconsistent with the hypothesis that S1 directly relays peripheral inputs, given that the observed cortical responses did not reflect the relatively constant activation over time expected to occur in the periphery in response to the simple PNS paradigm delivered to the nerve. If the cortex did directly mirror peripheral inputs, we would have expected that most electrodes would exhibit activation in the onset and sustained epochs (Figure 4A, green), rather than in onset only (Figure 4A, blue). Thus, our data suggests that the firing patterns induced in cortical neurons in response to PNS do not match a simple aggregation of PNS-elicited activation patterns in peripheral afferents (Figure 3B).

### Number of responsive channels increased as pulse width increased

As PW increased over the suprathreshold range, the total number of responsive electrodes increased in 9/10 sessions (slope line test, p < 0.01) (Figure 4C). Suprathreshold PW conditions (those above 0% PW) had a significantly greater responsive electrode count than the subthreshold -40% and -60% PW condition (Kruskal-Wallis test, p < .0001, post-hoc comparisons with Dunn-Sidak test, p < 0.04). However, there were no significant differences among the suprathreshold conditions. The positive relationship between PW and the number of responsive electrodes held for individual epochs of the cortical response as well as for the overall response. In other words, the number of electrodes with significant responses in the onset epoch significantly increased with PW, as did the number of electrodes with significant responses in the sustained epoch and offset epoch (Wilcoxon signed-rank test, p < 0.01) (*Figure 4*C, inset). The median slope of this relationship for electrodes responsive in the onset epoch was significantly greater than the median slope in the sustained epoch (Kruskal-Wallis test, p < 0.03, Dunn-Sidak post-hoc comparisons, p < 0.03) (*Figure 4*C, inset). Therefore, while the number of electrodes responsive to PNS increased with PW for all epochs, the number of electrodes that were responsive during onset increased with PW to a greater degree than the number of electrodes responsive in the sustained or offset epochs.

This finding could have resulted from two potential scenarios: 1) electrodes that were not responsive at low PWs became responsive at high PWs, and these new electrodes tended to include the onset epoch, or 2) electrodes that were responsive at low PWs changed their firing patterns to favor the onset transient at high PWs. To distinguish between these two scenarios, the behavior of the set of electrodes that were active at the 100% PW condition was examined across all PW conditions within each session. Overall, most electrodes retained the same pattern of responsivity across all PW conditions (*Figure 4*D, dark purple). In other words, if an electrode was responsive during the onset epoch at low PW levels, it would retain onset epoch responsivity as PW increased. In instances where electrodes changed their behavior with increasing PW, the change was most frequently adding additional epochs of responsivity (*Figure 4*D, light purple). For example, if an electrode was responsive during the onset epoch at low PW levels, it could become active during both the onset and offset epochs at higher PWs. This means that compared to the responsivity profile at 100% PW, the response at lower PWs would be some subset of the set of responsive epochs at 100%. Note that since the number of active electrodes tended to increase with PW (*Figure 4*C), many of the electrodes which exhibited responsivity at 100% PW were unresponsive at lower PW conditions (*Figure 4*D, light grey). These results are consistent with scenario #1, where more electrodes became responsive as PW increased, and their activity tended to include onset activity.

### Firing rate of individual channels increased as pulse width increased

We further characterized the magnitude and latency of the cortical activity during the onset and offset epochs and examined the relationships of these metrics with stimulation PW. The peak amplitude of the normalized firing rate in the onset epoch increased as the PW increased (linear regression t-test, p < .0001) (Figure 5A) and was statistically higher at successively higher PWs for all suprathreshold conditions except between 80% and 100% PW (Kruskal-Wallis test, p < 0.0001, with Dunn-Sidak post-hoc comparisons, p < 0.01) (Figure 5A). Latency, or the time from the start of stimulation to the peak amplitude in the onset response, was 177 ms +/- 89 ms on average across the suprathreshold conditions. Latency measures were normalized to the mean latency per session to determine the relative change in latency. Normalized latency for the onset epoch decreased as PW increased (linear regression t-test, p < 0.0001) (Figure 5B). On average, there was a 40 ms decrease in the time to reach the peak amplitude at the maximum PW (100% PW) compared to threshold (0% PW).

**Figure 5.**
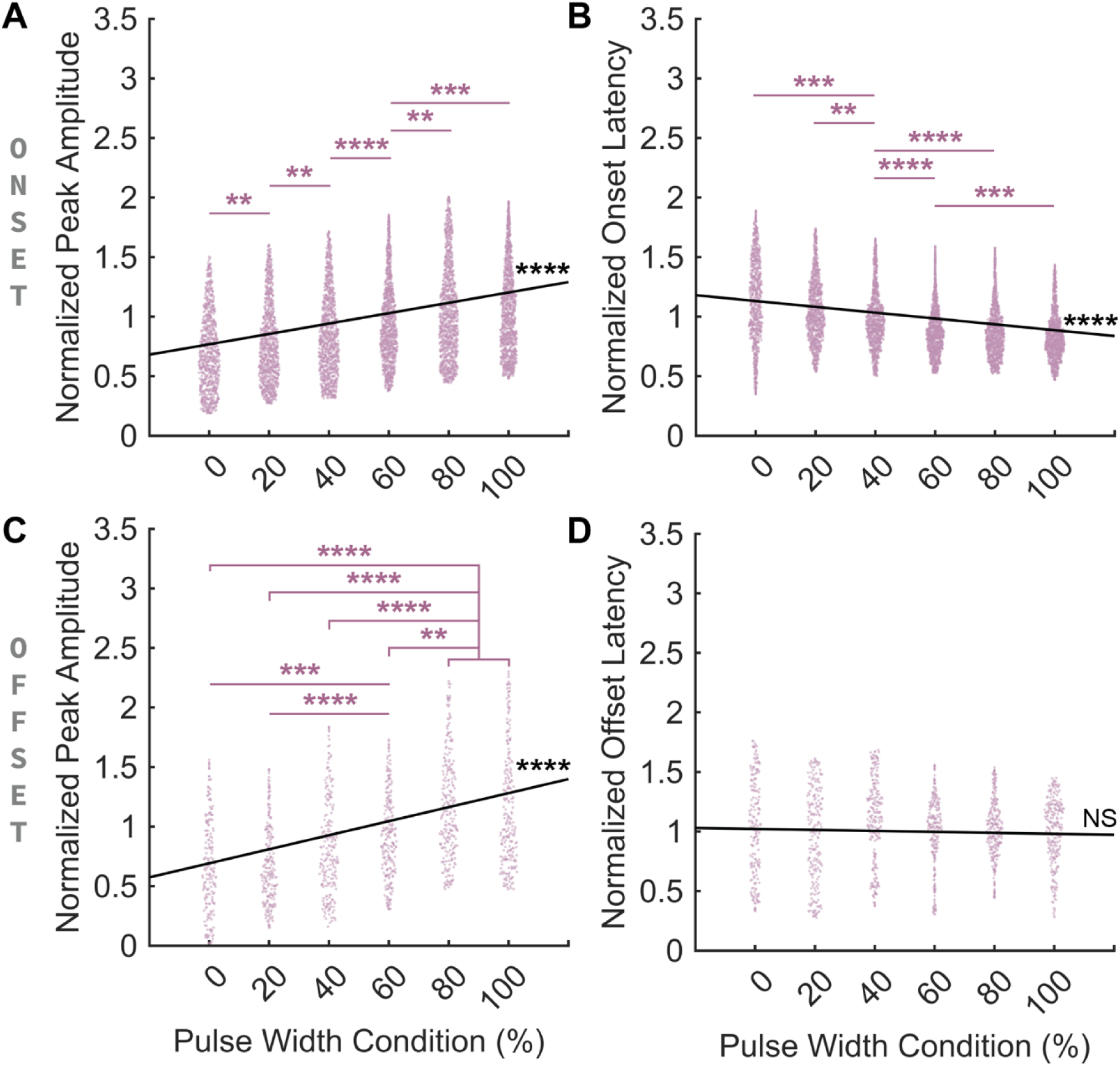
Peak amplitude and latency of the area 1 cortical response during onset and offset epochs for suprathreshold PNS conditions. A) Average normalized peak amplitude during the onset epoch aggregated across all sessions (n = 10 sessions, 1015-1384 points per PW condition, 7456 points total). B) Average normalized latency from stimulation start to the peak amplitude during the onset epoch, aggregated across all sessions (n = 10 sessions, 1015-1384 points per PW condition, 7456 points total). C) Average normalized peak amplitude during the offset epoch aggregated across all sessions (n = 10 sessions, 241-301 points per PW condition, 1657 points total). D) Average normalized latency from stimulation end to the peak amplitude during the offset epoch, aggregated across all sessions (n = 10 sessions, 241-301 points per PW condition, 1657 points total). For all panels: Purple asterisks denote significant differences between PW conditions, where ‘*’ indicates p < 0.05, ‘**’ indicates p < 0.01, ‘***’ indicates p < 0.001, and ‘****’ indicates p < 0.0001. The black line is a linear fit to the dataset presented within the panel. Black ‘****’ indicates a statistically significant slope of the relationship at p < 0.0001. ‘NS’ stands for not significant.

Similar to the onset epoch, the peak amplitude during the offset epoch also increased with stimulation PW (linear regression t-test, p < .0001) (Figure 5C). The amplitude of the offset response at 80% and 100% PW was significantly greater than all lower suprathreshold PW conditions and the amplitude at 60% PW was distinctly higher than 0% and 20% PW (Kruskal-Wallis test, p < 0.0001, with Dunn-Sidak post-hoc comparisons, p < 0.002) (Figure 5C). The average latency from stimulation end to the peak amplitude during the offset epoch across all suprathreshold conditions was 214 ms +/- 101 ms. Unlike onset latency, normalized latency to peak offset had no significant correlation to PW (linear regression t-test, p = 0.44) (Figure 5D).

### Different spatial activation in the cortex for different peripheral inputs

To understand the spatial distribution of cortical responses to PNS across the cortical arrays, heatmaps of the mean normalized activity during onset were generated per session and for each PW condition (Figure 6A). On average, the medial array had 6.7 times more activation during the maximum PW condition (100% of the sensory dynamic range) than the lateral array (Supplemental Figure 4). Due to the lateral array’s low activation across conditions and sessions, analyses of the location of cortical activation were restricted to the medial array.

**Figure 6.**
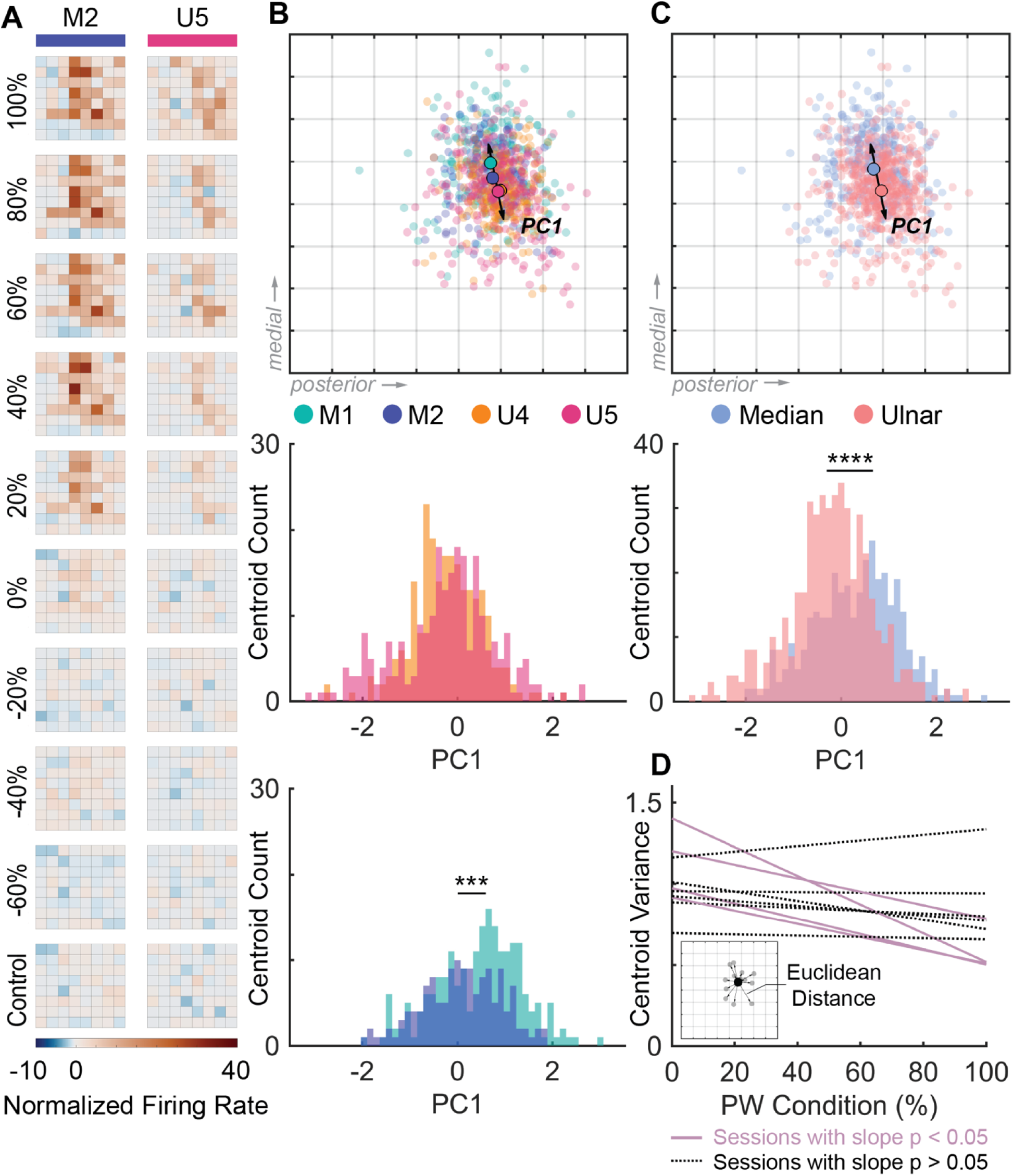
Spatial configuration of the cortical response to PNS. A) Normalized firing rate for each electrode in the medial S1 array during the onset epoch for each PW condition (rows) for a representative session of median nerve PNS (PNS of M2, left) and ulnar nerve PNS (PNS of U5, right). Each square in the grid represents the average firing rate relative to baseline during onset for a single electrode in the array across all trials for a given condition (n = 15 trials per condition for M2, 12-15 trials per condition for U5). B) Top: Weighted centroids of the location of the cortical response to PNS delivered through each C-FINE stimulation contact (M1, M2, U4, U5). Each small translucent point represents the weighted centroid for a single suprathreshold trial (n = 835 suprathreshold trials across all contacts and sessions). The larger points are the weighted centroids averaged across all suprathreshold conditions for each stimulation contact. Note that the average weighted centroid for U5 (pink) partially occludes the average weighted centroid for U4 (orange). The arrow represents the primary principal component (PC1) from the principal component analysis (PCA) used to reduce the 2D point space to 1D. The orientation of the array relative to the brain is marked with grey arrows on the right bottom region of the array. Middle: Histogram of the weighted centroid positions along PC1 per trial for U4 and U5. Bottom: Histogram of the weighted centroid positions along PC1 per trial for M1 and M2. ‘***’ denote p < 0.001. C) Top: Weighted centroids of the cortical response to PNS delivered to each nerve (median = blue, ulnar = coral). Small translucent points are individual trials and large points are averages for each nerve across all trials and sessions (n = 835 suprathreshold trials across both nerves). The arrow represents the primary principal component (PC1). Bottom: Histogram of the weighted centroid positions along PC1 per trial for each nerve. ‘****’ denote p < 0.0001. D) Variance in weighted centroid position across PW conditions. Each line (n = 10 sessions) represents the simple linear regression between the variance of the weighted centroid points across the suprathreshold PW conditions. The variance in threshold position significantly decreased with PW condition for four sessions (lavender lines) but did not significantly change in six sessions (dashed black lines). Inset: The variance in centroid position was calculated as the Euclidean distance between the average weighted centroid for a condition (black dot) and individual weighted centroids per trial corresponding to that condition (grey dots). In this example, there are 12 weighted centroids for the 12 trials with the condition, and hence 12 Euclidean distances that would be used to find the spatial variance.

In line with our previous findings, increasing PW led to more activity across electrodes (Figure 6A). However, the centroid of the spatial activity did not change across suprathreshold PW conditions (Figure 6A) (Kruskal-Wallis test, p > 0.13). Based on the average positions of each trial’s weighted centroid across all PW conditions and sessions, the center of activation was in the top right quadrant or the most posterior section of the array, regardless of which C-FINE contact was stimulated (M1, M2, U4, U5) (Figure 6B top). Within a nerve, the distributions of the center of activation were significantly different between the two median contacts (Rank-sum test, p < .0002) but were not significantly different between the two ulnar contacts (Rank-sum test, p = .51) (Figure 6B bottom). When comparing the center of activation across C-FINE contacts between the two nerves (Figure 6C top), the distributions of the center of activation were significantly different in the medial-lateral direction for median vs. ulnar PNS (Rank-sum test, p < 0.0001) (Figure 6C bottom).

Next, we examined the variability in the center of activation across trials for each suprathreshold PW to understand the consistency of the cortical hotspot responding to the stimulation. Variability of the centroid across trials was measured as the Euclidean Distance (ED) between each trial’s weighted centroid and the average weighted centroid for the set of trials that share the same PW condition (Figure 6D left). In most sessions (6/10) there was no change in the weighted centroid with changes in PW. In 4/10 sessions, the variability of the weighted centroid decreased as PW increased (linear regression t-test, p < 0.03) (Figure 6D right). Stated another way, at high PWs, the center of activation varied over a smaller area than at lower PWs in these sessions.

### Onset response has the strongest correlation to perceived intensity

Finally, we investigated which epochs in the cortical activity resulting from PNS best predicted the perceived intensity ratings reported by the participant. For this analysis, we considered the aggregate activity of the entire recorded neuronal population, because we wished to understand how the behavior and computations across the ensemble of neurons reflected the intensity of perception^36^.

We performed principal component analyses (PCA) of the average normalized firing rates for each epoch (i.e., onset, sustained, and offset) across all the 128 S1 electrodes on a per session basis. For each of the three epochs, we then found the set of principal components (PCs) per session that cumulatively captured 60% of the variance of the activity in that epoch. The number of principal components needed for capturing 60% of the total variance during the onset epoch was significantly greater than the number required for the sustained epoch (Kruskal-Wallis test, p < 0.05, with Dunn-Sidak post-hoc comparisons, p < 0.05), suggesting that the represented sensory information is more distributed across a higher dimensional subspace during onset compared to the other epochs.

We then constructed three multiple regression models per session – one regression model for each of the three epochs (i.e., onset, sustained, offset) – that predicted perceived intensity from these down-selected sets of principal components (Figure 7A). The onset models demonstrated moderate but significant fits to the perceived intensity ratings (average adjusted R^2^ of 0.24 +/- 0.13 across sessions), while the sustained and offset models generally had poor fits (average adjusted R^2^ of 0.04 +/- 0.08 for sustained model and adjusted R^2^ of 0.05 +/- 0.13 for offset model across sessions) (Figure 7B). Furthermore, only the onset models exhibited adjusted R^2^ values that were significantly above chance (Wilcoxon signed-rank test, p < 0.002) (Figure 7B). This demonstrates that the information contained in the cortical activity during the onset epoch is most strongly related to the perceived intensity ratings, compared to the cortical activity during the other epochs. Additionally, the onset models had a greater percentage of predictors that were significant, compared to the sustained models (Kruskal-Wallis test, p < 0.02, with Dunn-Sidak post-hoc comparisons, p < 0.02) (Figure 7C). In fact, the sustained and offset models for most sessions had no significant predictors (Figure 7C, scatter points at zero). Across all models, the principal components that explained the most variance (i.e., PC1, PC2, etc.) were not consistently those that significantly predicted perceived intensity (Supplemental Figure 5). In addition, the principal components that significantly predicted perceived intensity were not consistent across the regression models (Supplemental Figure 5).

**Figure 7.**
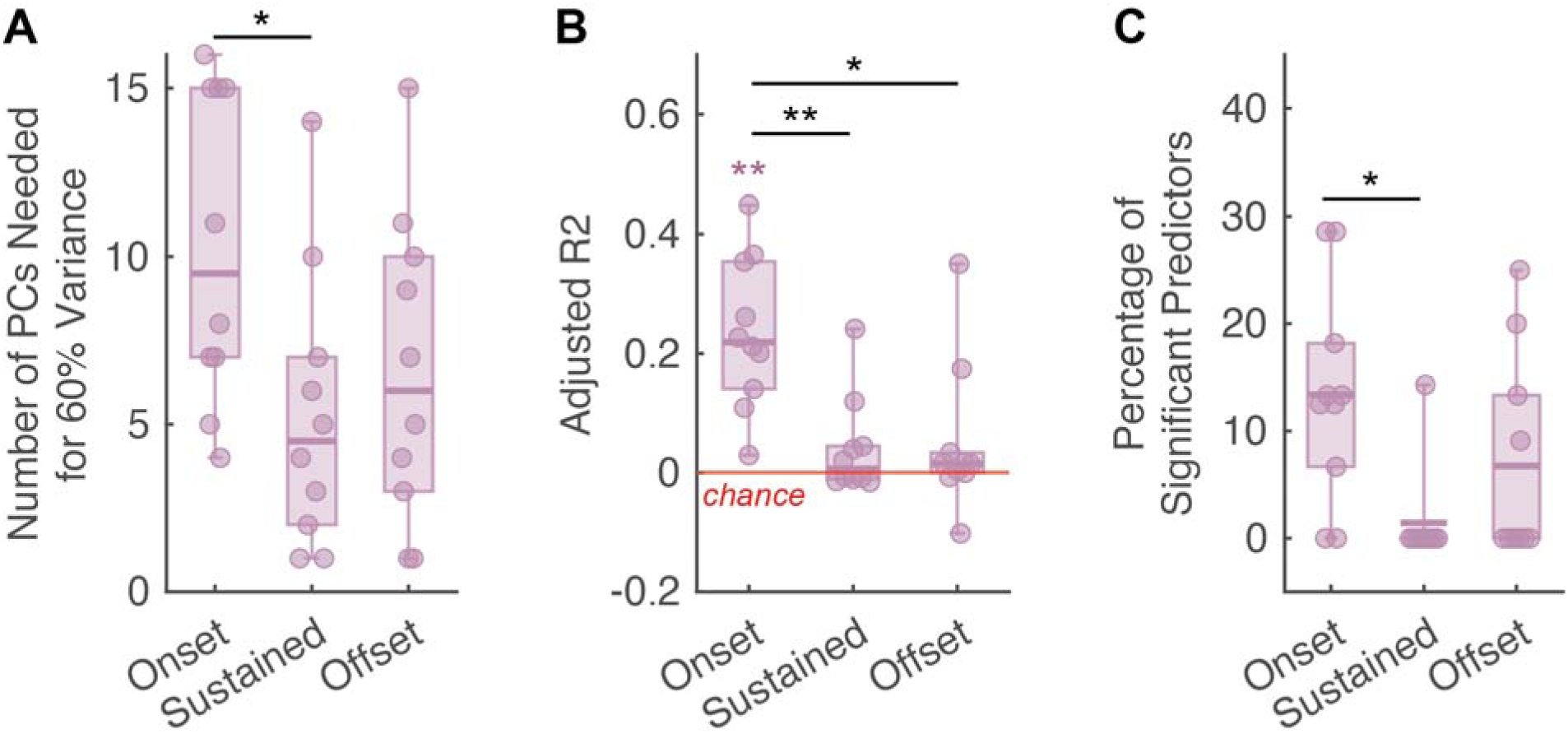
Multiple linear regression models predicted perceived intensity ratings from the cortical population activity in the onset, sustained, or offset epochs. A) The number of principal components that account for 60% of the total variance for each epoch, calculated from a principal component analysis on the activity across the 128 S1 electrodes. Separate PCAs were run for each epoch and session (n = 10 sessions). B) Adjusted R^2^ values for regression models relating population activity in the onset, sustained, or offset epochs to perceived intensity. Separate models were fit for each epoch and session (n = 10 sessions). Each regression model consisted of predictors which were the set of principal components required to account for 60% of the variance in the given epoch’s dataset. The red line is the adjusted R^2^ value at chance. Purple ‘*’ denotes adjusted R^2^ values that are significantly different from chance. Black ‘*’ shows significant differences in adjusted R^2^ values across epochs. C) Percentage of PCs included in each regression model that were significant predictors of perceived intensity (n = 10 sessions). For all panels, ‘*’ signifies a difference with p < 0.05 and ‘**’ signifies a difference with p < 0.01.

## Discussion

### PNS is an effective approach to provide sensation to people with sensory-incomplete SCI

Despite having a sensory-incomplete SCI, the participant reliably perceived PNS delivered below-injury. The participant perceived sensation from PNS on regions of the hand matching the innervation territory of the stimulated nerve, felt several distinct levels of intensity with increases in PW, and reported quality descriptors similar to those reported in prior studies of PNS in individuals with limb loss^19,26^.This below-injury sensation from PNS provides evidence that at least some of the connections between the first-order sensory afferents and the dorsal column-medial lemniscal (DCML) pathway were preserved. Our findings about the perceptibility of PNS demonstrate that there is value in pursuing integration of PNS as a sensory feedback modality for bidirectional neuroprostheses. PNS could be used to supplement the users’ existing touch capabilities, which may have been damaged by the SCI, to ensure that they can reliably detect sensation and feel distinct levels of tactile intensity. PNS may provide a more robust signal to the brain than the user’s natural touch can provide, because PNS can provide consistent input regardless of the contact area or dynamics of the physical object interaction and can scale the sensation over a broader range by recruiting large afferent populations - up to one or more fascicles - at once. PNS may also offer different qualia, such as vibration, that are not readily perceived through the participant’s residual tactile pathways^37^. Additional qualia could provide more enriching information when interacting with their surroundings using assistive devices or motor neuroprostheses, which may improve their ability to make decisions and respond appropriately during complex control tasks.

In addition to perceiving suprathreshold PNS with high accuracy, the participant occasionally reported perceiving subthreshold PNS, as indicated by non-zero intensity ratings for some subthreshold stimuli. The occasional non-zero intensity values on subthreshold conditions (Figure 2C) may indicate participant bias toward reporting perception or may have arisen due to changes in threshold that caused the previously subthreshold stimuli to become suprathreshold. Supporting the latter explanation, we observed progressive decreases in sensory thresholds from block to block in most sessions. While the reason for the decreasing sensory thresholds is unclear, we anecdotally observed that the decreases were most likely to occur after large SCI-related spasms. These spasms have occurred sporadically since the participant’s injury, including on days without PNS application, and thus do not appear to be caused by PNS. To address the threshold instability issue, we repeated the thresholding procedure and recalibrated the stimulation levels between each block to ensure consistency in the sensory dynamic range throughout the session.

### Cortical responses to PNS suggest central mechanisms of cortical onset transients

As expected, PNS levels that generated perceivable sensation also produced cortical activity in S1 that was distinguishable from baseline activity. Notably, the cortical responses to PNS typically had strong onset transients. This finding is not consistent with a paradigm where cortical activity patterns result from directly relaying and/or aggregating peripheral afferent activation patterns. The existing evidence suggests that PNS induces synchronous activation of a mixed population of peripheral nerve RA and SA afferents throughout the entire PNS stimulus^31^. If the cortical activity from PNS had matched the peripheral activation pattern, we would have expected that most cortical electrodes would display above-baseline activation throughout the entire PNS stimulus. Instead, the top three most prevalent firing patterns we observed in the S1 cortical response to PNS were above-baseline activation in the onset period only, in both the onset and offset periods, and in the offset period only (Figure 4A). These response patterns to PNS resemble the “RA-like” firing patterns observed in S1 following mechanical indentation of the fingers, with elevated firing at the onset and offset of the mechanical stimulus^32,33,35^. Our finding that the predominant response profile in S1 was activation solely during onset also mirrors the patterns of cortical evoked potentials found in a prior NHP study for both constant frequency PNS and vibrotactile stimuli^33^. Another NHP study of vibrotactile stimulation showed a strong onset transient and a much less pronounced sustained response in S1^38^, which also bears similarity to our observed S1 responses to PNS. Therefore, while the cortical responses to PNS do not directly match the expected peripheral responses to PNS, the temporal firing patterns in the cortex are qualitatively similar to RA-like behavior from mechanical stimuli, including both indentations and vibrotactile stimulation.

One possible reason why the cortical response to PNS is different from the peripheral response and instead displays a strong onset transient is because of sub-cortical processing in the cuneate nucleus (CN) of the thalamus. A prior NHP study that examined the CN response to mechanical indentation showed that the CN neurons exhibit submodality convergence and that the majority of their responses tended to be more RA-like than SA-like^39^. Interestingly, the percentage of RA-like responses in the NHP CN was greater than the percentage of RA responses in the periphery^39^. Similarly, in the cortex, the percentage of RA-like responses was far greater than SA-like responses, and these RA-like responsivity percentages were higher in area 1 of S1 than in area 3b^32^. Taken together, this evidence suggests that the input from RA fibers is being amplified as the signal travels up the DCML pathway. The cortical response to PNS observed in this study further supports the idea that the peripheral signal is being transformed to emphasize the onset transient as it travels up the DCML pathway.

Moreover, our findings allow us to disambiguate whether the pronounced RA-like response in area 1 arises directly from the peripheral afferents in the nerve or arises from the intervening subcortical and cortical circuits (e.g., CN, area 3b, and/or area 1 itself). With mechanical touch inputs, it is impossible to distinguish between these two scenarios, since there are more RA than SA fibers in the periphery, and peripheral RA fibers innately display distinct onset transients. Thus, RA-like response profiles at the cortical level could result from preferentially relaying RA inputs from the periphery along the DCML pathway or could reflect local circuit dynamics that produce cortical onset transients. Since PNS is theoretically forcing both peripheral RA and peripheral SA fibers to fire throughout the entire stimulation duration, but area 1 neurons are still displaying a strong onset transient reminiscent of RA-like activity from mechanical touch, this study provides evidence that cortical neuron firing patterns do not result from simply relaying peripheral firing patterns. Instead, subcortical and cortical circuits play a role in producing onset transients.

### Cortical activity reflects changes in PNS input

We found that the number of responsive electrodes, the peak activation amplitude during stimulus onset and offset, and the latency of the peak amplitude modulated with increases in PNS PW. The increase in the responsive electrode count suggests that more cortical neurons are responding to PNS as PW increases. Given that increasing PW of PNS is known to recruit a larger fiber population in the periphery^40^, this indicates that larger activated peripheral afferent populations lead to larger activated cortical populations. The increase in peak amplitude during stimulation onset and offset with increasing PW means that responsive cortical neurons also increase their firing rate with increases in PW. These findings are consistent with a prior study showing how increasing indentation depth on the fingertips increased the firing rate of individual S1 neurons as well as the spread of activation in S1^35^. The increase in activated peripheral afferent population size due to increases in PW may also be responsible for the decrease in latency between stimulation onset and the peak amplitude during the onset period observed in our data at higher PWs. More recruited peripheral afferents could allow faster action potential generation at downstream synapses because spatial summation of more incoming signals would enable the post-synaptic neurons to surpass firing threshold more easily^41^, leading to faster transmission along the DCML pathway and an overall decrease in latency at the cortex.

We also found that all PNS stimuli, regardless of the nerve or fascicle activated, resulted in cortical activation in a similar area of the medial array. This finding suggests that this specific part of the S1 arrays predominantly captured activity from the PNS inputs, likely because it had the best placement of electrode contacts within the relevant cortical layers^11,42^. We previously reported that delivering intracortical microstimulation to this region of the medial array generated more sensory percepts relative to other regions of the array^11^, which further supports the electrode placement hypothesis. Our data also shows that a relatively small cortical region, captured by cortical recordings from a few S1 electrodes in the centroid region, can respond to peripheral inputs from both the median and ulnar nerves. Thus, relatively small regions of S1 can represent touch stimuli spanning the entire anterior surface of the hand, providing further evidence that the receptive fields of afferents in S1 are larger than their projected fields might suggest^8,11^. The fact that the weighted centroid location does not move with increasing PW suggests that the additional recruited peripheral fibers all have connections in approximately the same cortical region. This potentially could suggest that the additional afferents recruited in the periphery from increasing PW are contained within a single fascicle that has a relatively small receptive field. We also speculate that the trend of decreasing trial-to-trial variability of the centroid with increasing PW was caused by the activation of the same or closely located S1 electrodes across a greater number of trials as PW increased.

### Cortical biomarkers of touch perception

One goal of this study is to identify cortical biomarkers that correspond to aspects of touch perception. Our analysis demonstrated that cortical recordings in S1 display markers for the start and end of PNS, as evidenced by the strong onset and offset transients in the cortical signals. Since PW correlates to the number of active electrodes and the magnitude of the onset response, and perceived intensity scales with PW, we can infer that perceived intensity would have the same correlation to the cortical biomarkers as we showed PW to have. We further sought to understand how well the array-wide responses in different epochs correlated with perceived intensity ratings using multivariate linear regression. Models based on the onset epoch had the best fit to perceived intensity, indicating that the activity of S1 neurons was most correlated with perceived intensity in the first several hundred milliseconds of their response. Although there may have been modulation in S1 during the sustained and offset epochs, it was not as strongly related to perceived intensity as was the activity in the onset epoch. Therefore, onset activity across the activated population of cortical neurons may be a useful biomarker for the perceived intensity of the evoked sensation.

However, while the firing rate during the onset epoch predicted perceived intensity better than firing rates in any other epoch (average adjusted R^2^ of 0.24 +/- 0.13 for the onset models), a simple linear regression based on suprathreshold PW far outperformed all of the cortical metrics in predicting perceived intensity (average adjusted R2 = 0.81 +/- 0.07). This is likely because the participant was highly consistent in the perceived intensity ratings he gave for each PW condition, whereas there was much higher variability in the cortical population activity across trials, which likely contributed to the poorer model fits for models based on cortical activity. Thus, better signal quality and larger datasets would aid future investigations into cortical biomarkers of perceptual experiences. In addition, a limitation to our regression analyses is that the predictors in these regression models were the cortical data projected onto the set of PCs that cumulatively accounted for 60% of the total variance. This approach assumed that the variance in the data was primarily related to the perceived intensity. However, if the majority of variance was not linked to intensity and was rather capturing changes in attention, noise, or some other factor, our approach to building the regression models based on the top PCs would have yielded poor model fits and few predictors that were significantly related to perceived intensity. This likely occurred in some sessions for the sustained and offset models, which had low adjusted R^2^ values not distinguishable from chance and few PCs that were significant contributors to the model.

Additionally, our study did not attempt to provide any new insight on the cortical biomarkers of sensation quality. Most of the sensory percepts evoked by PNS in this study were described by the participant as tingling or paresthesia. This type of sensation does not match the perceived quality of mechanical touch applied to the hand, which the participant has typically described as pressure or touch^43^. Since both PNS and mechanical touch of the hand are delivered below injury, we would expect the SCI to influence them both similarly. Despite the distinct differences in percept quality between PNS and mechanical touch, the S1 responses to PNS qualitatively looked similar to those expected to arise from mechanical indentation, at least in the single channel analyses predominantly performed in this study. While it is possible that there are differences in other single channel metrics or population-wide cortical metrics between PNS and mechanical stimuli that may explain their perceptual differences, our experiments were not set up to investigate this question, as we did not deliver or assess mechanical stimuli during these experimental sessions. In addition, we also could not explain differences in perceived quality across PW conditions because we did not collect sensation quality on a trial-by-trial basis. A direct comparison study between PNS and mechanical touch that involves both single channel and population-wide cortical metrics may reveal biomarkers of sensory quality, since quality perception may be explained via more complex neural circuits integrating various brain regions^50^.

### Implications for biomimetic stimulation approaches

We designed this study to understand the S1 response to the most basic PNS paradigm to serve as a foundation for future studies investigating more complex PNS paradigms. The PNS stimuli in this study were simple pulse trains lacking any spatiotemporal parameter variations. However, recent studies have developed biomimetic PNS paradigms, which replicate the temporal profile of the peripheral response to natural touch by delivering a higher amplitude or frequency at the onset and offset of the pulse train^44,45^. Preliminary studies implementing such biomimetic patterns demonstrated that they could increase the naturalness of sensation^45^. A prior NHP study also showed that a biomimetic PNS paradigm produced cortical evoked potentials that had increased firing at the onset and offset of the stimulus, similar to the pattern resulting from mechanical indentation^33,46^. In future work, we will examine S1 responses in humans to biomimetic PNS and other spatiotemporal PNS paradigms and develop more complex cortical metrics to enable systematic comparisons among PNS paradigms as well as between PNS and mechanical touch inputs. Conducting these studies in humans will be critical to understanding the relationships between cortical activity and aspects of the perceptual experience of PNS, which will support the development of enhanced PNS approaches for restoring function to people with neurological disorders.

## Data Availability

All data produced in the present study are available upon reasonable request to the authors.

## Acknowledgments

We thank our participant for his dedication to the study and for all the time he has volunteered, as well as his family for supporting him throughout the clinical trial.

## Author Contributions

E.L.G and P.B. designed the study. P.B. performed data collection and analysis. W.D.M. and A.K.O. provided engineering support. B.H. and B.S. designed aspects of experimental software. C.F., D.T., R.B., and B.H. aided with neural data post-processing and analysis techniques. P.B., E.L.G., and A.B.A. interpreted the results. P.B. wrote the initial paper draft and all authors provided critical review, edits, and approval for the final manuscript. J.P.M., J.A.S., and E.H. performed surgical implantation procedures and managed medical care of the participant. E.L.G., A.B.A., and R.F.K. managed oversight for the clinical trial. E.L.G. supervised the study.

## Materials and Methods

### Participant and Surgery

This study was performed with a male participant with C3/C4 tetraplegia recruited to the Reconnecting the Hand and Arm to the Brain (ReHAB) clinical trial when he was in his late twenties (ClinicalTrials.gov ID: NCT03898804). His motor-complete, sensory-incomplete injury is categorized as Grade B on the American Spinal Injury Association Impairment Scale (AIS-B).

The cortical implant was comprised of six 8×8 (64-electrode) iridium oxide microelectrode arrays (MEAs) (Blackrock Neurotech, Salt Lake City, UT) placed in regions of the left hemisphere involved in upper extremity function, including two MEAs in S1. MEA locations in S1 were determined using awake stimulation mapping with an Ojemann stimulator (Integra Lifesciences, Plainsboro Township, NJ), which applied stimulation to the cortical surface. The lateral (electrodes 1-64) and medial (electrodes 65-128) arrays were placed over S1 regions eliciting sensation on the ring and index fingers, respectively, during intraoperative testing^11^. In a separate surgery, nine 16-channel composite flat interface nerve electrodes (C-FINEs) (Ardiem Medical, Indiana, PA) were placed around the nerves innervating the arm and hand, including the median and ulnar nerves^11^. For more information about the implanted devices and surgical approach, please see Herring et al. 2023^11^. To assess his residual sensory capabilities, the static two-point discrimination test^46^ and the Semmes-Weinstein monofilament test^47^ were performed prior to his implant and then reassessed after his implant. Near-normal values were found on all fingers excluding the thumb, which had more reduced values (Supplemental Table 1). There were no significant changes to his residual sensory capabilities after the implantation surgery.

All procedures were reviewed and approved by the University Hospitals Cleveland Medical Center’s Institutional Review Board. The U. S. Food and Drug Administration also approved the devices and procedures for this study under an Investigational Device Exemption. The participant gave informed consent for study enrollment before the implantation procedure and any experimental sessions. Sessions to collect data for the experiments described in this manuscript occurred between months 10 and 37 after implant. A total of 14 sessions were conducted. Cortical data was usable from 10 of these sessions, while all 14 sessions were included in the perceptual analyses (Supplemental Table 2).

### Peripheral Nerve Stimulation

Current was delivered from individual C-FINE contacts via an external, current-controlled stimulator known as the Universal External Control Unit (UECU) (Technical Development Laboratories, Case Western Reserve University, Cleveland, OH). The return path for the stimulation current was a single large C-FINE contact that is eight times the area of the other 15 C-FINE contacts. The charge density per phase and charge per phase of the stimulation pulses never exceeded the Shannon curve limits for safe stimulation of tissue^48^. The stimulator could deliver current in increments of 0.1 mA between 0.1-2 mA. PW could be delivered between 1-255 us in 1 us increments. All pulse trains were comprised of charge-balanced, square-wave, biphasic pulses with the cathodal phase first. Pulse frequency (PF) was fixed at 50 Hz for all experiments in the study.

### C-FINE Characterization and Thresholding

Initial thresholds at the far right of the strength-duration curve were found after implantation using an ascending staircase procedure. To find the pulse amplitude (PA) threshold, PF was fixed at 50 Hz and PW was fixed at 250 us. The PA was increased in 0.1 mA increments until the participant perceived the stimulus. Next, the PA was fixed at this threshold value, and a binary search was conducted to find the PW threshold. This thresholding procedure was performed on each C-FINE contact around the median and ulnar nerves. The projected fields at threshold for each of these contacts can be viewed in Supplemental Figure 1. Four of the contacts that elicited sensory percepts (M1 and M2 contacts on median, U4 and U5 contacts on ulnar) were selected for further testing. The experiment was run on each of these contacts between 2-3 times, except for U5, which was tested 6 times to replace sessions with poor signal quality (see below for exclusion criteria).

### Establishing Conditions

For each session, one C-FINE contact that had previously been found to produce sensation was chosen for stimulation delivery during the experiment. The PA on that contact was set to the value found initially at sensory threshold, and an ascending staircase approach was used to re-find the PW corresponding to detection threshold. Next, the PW was gradually increased in 10-20 µs increments until the participant reported experiencing the sensation at their maximum comfortable level. Based on this mapping procedure, we defined the sensory dynamic range as the range of PW values spanning from detection threshold to the maximum comfortable level. PW conditions were then labelled as some percentage of the sensory dynamic range, with 0% being sensory threshold and 100% being maximally comfortable. There were a total of ten PW conditions: six PW values that were equally spaced across the suprathreshold sensory dynamic range (0%, 20%, 40%, 60%, 80%, 100%), three equally spaced PW levels that were subthreshold (-20%, -40%, -60%), and a control condition of no stimulation. Each session consisted of 3-8 blocks of 30 trials each, resulting in 9-24 repetitions per stimulation level. If the sensory threshold appeared to change between blocks within a session, we repeated the thresholding procedure described earlier to find the new PW values at threshold and maximum comfortable, and recalculated the PW values for all conditions relative to the new sensory dynamic range before continuing onto the next block of the experiment.

### Experimental Trial Structure and Intensity Estimation

Each trial consisted of a PNS pulse train presented for two seconds. The ten selected PW conditions were pseudorandomly varied across trials within each block of the session. The experiment was single-blinded for PW condition. For each trial, the participant was asked to report the perceived intensity of the PNS-evoked sensation on an open-ended scale of their choosing. The participant was instructed that if the evoked percept felt twice as intense as the previous trial, they should give it a rating that is twice as large as the rating ascribed to the previous trial. Similarly, if the sensation felt half as intense, the participant was asked to report a value half as large. If the stimulation was not perceived during a trial, they were instructed to assign a rating of zero.

### Perceptual Data Analysis

Perceived intensity ratings were normalized to the mean of each experimental block to allow for aggregation of data across blocks and sessions. The Gompertz function was used to create a sigmoidal fit between the PW condition (as a percentage of the sensory dynamic range) and the normalized intensity values. The relationship between the percentage of the sensory dynamic range and normalized intensity was found using a simple linear regression and the statistical significance of the slope was determined using the linear regression t-test. Normalized intensity distributions were also compared across different PW conditions with an ANOVA and Tukey-Kramer post-hoc comparisons. An ANOVA was also used to compare the normalized intensity distributions across stimulated contacts, corresponding nerves, and days when sessions occurred.

### Neural Feature Extraction and Data Cleaning

In parallel to collecting perceptual metrics, multi-unit activity from the two 64-channel MEAs in S1 were recorded using Cerebus Neural Signal Processors (Blackrock Neurotech, Salt Lake City, UT) at a rate of 30K samples/sec. This data was collected for twelve of the fourteen total sessions. Following analog and digital bandpass filtering from 250-5000 Hz and digital common average referencing, individual spike waveform occurrences (a.k.a. threshold crossings) were extracted using a -4.5σ threshold based on the root mean square noise of the filtered continuous data^49^. Threshold crossings occurring within a 6-second interval, spanning two seconds before, two seconds during, and two seconds after each PNS stimulus, were converted into firing rates using 1 ms time bins. This neural feature was then smoothed using a Gaussian kernel (s.d. of 50 ms) and normalized to baseline activity, defined as the activity occurring in the two second interval prior to stimulation onset for each trial. Specifically, the firing rate during each trial was z-scored using the following formula, which was modified from a prior similar study^35^: 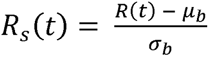, where *R(t)* is the firing rate at a given time bin *t*, and *µ* and *a*_b_ are the mean and standard deviation of the baseline firing rate averaged across all the trials of an experimental block. Peri-stimulus time histograms (PSTHs) of condition-averaged normalized firing rates per electrode were generated to visualize the firing patterns.

To minimize the effects of a poor connection to the array or array-wide noise, only trials and electrodes that demonstrated low noise and adequate neural firing were considered for further analysis. Trials were excluded if they a) had low firing rate (i.e., firing rate < one spike over the six-second trial interval) on 75% of the electrodes, b) had high firing rate (i.e., over 1000 Hz firing at any point throughout the six-second trial interval) on 75% of the electrodes, or c) had highly correlated firing rates across a majority of the electrodes. Specifically, correlation among electrodes was measured using Pearson’s coefficient between electrode pairs, and to be excluded, the trial had to have a Pearson’s coefficient value greater than 0.5 across 50% of unique electrode pairs. Because over 50% of trials were excluded for two sessions using these criteria, we excluded those sessions from further neural analysis (Supplemental Table 2). Since both of these excluded sessions involved PNS on contact U5, we collected two additional sessions with contact U5. We also excluded electrodes from the analysis on a per session basis. Electrodes were excluded from a given session’s analysis if 90% of trials in that session on that electrode were categorized as having low or high firing activity as defined above.

### Neural Data Analysis

Each trial was divided into four epochs for further analysis. The baseline epoch started 2000 ms prior to stimulation and lasted for 1600 ms, the onset epoch began at the start of the PNS pulse train and lasted for 400 ms, the sustained epoch began at the conclusion of the onset epoch and lasted for the remaining 1600 ms of the PNS pulse train, and the offset epoch began at the end of the PNS pulse train and lasted for 400 ms after stimulation ended. Our first objective was to determine which electrodes were responsive during these epochs of the trial. Therefore, for each of the 128 electrodes, the normalized firing rate was averaged across each of these epochs per trial. Per electrode, an ANOVA was performed on the mean normalized firing rate in each epoch across trials within a condition, followed by Dunnett’s test to compare the means during the onset, sustained, and offset epochs against the baseline. If there were at least two or more suprathreshold conditions where the mean activity in any epoch was greater than the mean activity during baseline, then the electrode was classified as having an ‘excitatory response’ at those PW conditions. Similarly, if there were at least two or more suprathreshold conditions where the mean activity in any epoch was less than the mean activity during baseline, then the electrode was classified as having an ‘inhibitory response’ at those PW conditions. The Wilcoxon rank sum test was used to compare the percentage of excitatory responses to the percentage of inhibitory responses across sessions. Only those electrodes responsive to PNS in at least one epoch with excitatory behavior in at least two suprathreshold PW conditions were included in the following analyses unless stated otherwise (varied from 1 to 31 electrodes per session).

After identifying the electrodes that were responsive to the stimulation, we sought to characterize their activity by defining their ‘activation profiles.’ These activation profiles were categorized by the epochs with activity significantly greater than baseline (ANOVA with Dunnett post-hoc comparisons) into one of seven patterns (see Figure 4A). An ‘activation profile’ is the trial-averaged, normalized firing rate trace for a PW condition where the electrode was responsive. For instance, if one electrode was responsive in some combination of epochs during three suprathreshold PW conditions and another electrode was responsive during only two conditions, this would result in a total of five activation profiles. The Kruskal-Wallis test with Dunn-Sidak post-hoc comparisons were used to compare the percentage of activation profiles in each of the seven patterns across sessions.

Next, we quantified the total number of electrodes responsive in either the onset, sustained, offset, or some combination of all three epochs for each PW condition per session. A simple linear regression was computed per session between the percentage of the sensory dynamic range and the responsive electrode count for the four cases. The Wilcoxon signed rank test was performed to see whether the slope groups per epoch were significantly greater than 0 (n = 10 sessions). The slope groups were also compared across individual epochs with the Kruskal-Wallis test using Dunn-Sidak post-hoc comparisons.

To examine how an electrode’s activation profile changed across PWs, we first found the set of electrodes responsive in one or more epochs at the 100% PW condition (n = 103 electrodes total across sessions). For each of these qualifying electrodes, the activation profiles for each lower PW condition were compared to the activation profile at 100% PW. If an electrode at the lower PW condition was responsive in the same epochs as at 100% PW, it was labeled as having the same responsivity. If it was responsive in only a subset of the epochs observed at 100% PW, it was labeled as having fewer epochs than 100% PW. If it was responsive in additional epochs beyond those at 100% PW, it was labeled as more epochs than 100% PW. If the electrode at the lower PW was responsive in entirely different epochs than at 100% PW, then it was classified as having a different responsivity pattern. Lastly, if there were no epochs that were responsive, the electrode was labeled as not responsive for that condition.

Then, we examined the peak amplitude of the normalized firing rate on a trial-by-trial basis during the onset and offset epochs for responsive electrodes at suprathreshold PWs (52-126 trials per session after trial exclusion outlined above). Peak amplitude at onset was defined as the maximum normalized firing rate within the first 400 ms following stimulation start, and peak amplitude at offset was defined as the maximum normalized firing rate within the first 400 ms following stimulation end. Latency was defined as the time it took to reach the peak amplitude in the onset and offset epochs from stimulation start and end, respectively. For onset, there were 1742-1810 data points for each PW condition, which arose from multiplying the trial count for a session with the number of responsive electrodes and aggregating across sessions. For offset, there were a total of 367-382 points per PW across sessions. If the latency for a trial on an electrode was found to be on either boundary of the epoch’s time window (e.g., 0 or 400 ms for the onset epoch), it indicated the absence of a peak within the epoch, since the likelihood of the normalized firing rate having a peak at exactly the epoch’s boundaries is extremely low. In these cases, we removed that trial from analysis, since the peak could not be reliably found. Consequently, approximately 30% of points across the suprathreshold conditions were removed from these analyses. To account for electrode-to-electrode variability in amplitude and latency, each electrode’s metrics for each trial were divided by the mean value on that electrode across trials to get the relative change of amplitude or latency. After this normalization technique, the metrics were aggregated across the 10 sessions. The slope of the relationship between the suprathreshold conditions and these metrics was assessed for statistical significance using the linear regression t-test. Additionally, the distributions for each of the metrics were compared across suprathreshold PW conditions using the Kruskal-Wallis test with Dunn-Sidak post-hoc comparisons.

The center of activation for both lateral and medial arrays was determined with a weighted centroid across the arrays for each suprathreshold trial (835 suprathreshold trials total across sessions). The weighted centroid was calculated by weighting each electrode by its average normalized firing rate during the onset epoch and taking the 2D average of the weighted centroid positions. The lateral array had an average of 6.7 times lower activation overall compared to the medial array at 100% PW across all sessions. In addition, the activity on the lateral array did not form a distinct cluster of activation, and thus the center of activation on the array could not be defined. Therefore, the lateral array was excluded from this analysis, and the following analyses were performed on the medial array only. Principal component analysis (PCA) was then used to reduce the 2D space of the weighted centroid points to 1D, enabling a statistical comparison of the distributions of the weighted centroids across different PNS inputs (Wilcoxon rank-sum test) and stimulation conditions (Kruskal-Wallis test). Variability of the weighted centroid points across trials for a given suprathreshold PW condition was quantified by calculating the Euclidean distance between the weighted centroid of an individual trial and the average weighted centroid for all trials at its corresponding PW condition. On a per session basis, a simple linear regression between the suprathreshold PW conditions and this Euclidean distance variability metric was performed, and the statistical significance of the regression slope was found. The Euclidean distances were also compared across suprathreshold PW conditions with the Kruskal-Wallis test.

Lastly, we studied the relationship between population cortical activity during different epochs of the stimulus and perceived intensity ratings on a per-session basis using multivariate linear regression. To do this, we first found the normalized firing rate averaged during the onset, sustained, and offset epochs for each trial across each of the 128 electrodes per session. Only trials at threshold and above (0% - 100% of the sensory dynamic range) that were perceived were retained for this analysis. Separate principal component analyses (PCA) were performed on the activity averaged for each of the three epochs of interest across all electrodes. Each trial’s firing rate data for each epoch was then projected onto the principal component axes found for that epoch to represent the population activity across channels in a reduced dimensional space. For each epoch, these transformed principal components then served as the set of predictors for a multivariate linear regression to predict perceived intensity. The number of predictors for each epoch’s multivariate linear regression was the set of principal components needed to cumulatively account for 60% of the variance in that epoch across trials. We then ran these multivariate regression models per epoch and session. Adjusted R^2^ was used to compare how well the models from each epoch related to the perceived intensity. Adjusted R2 was used because it considers the number of predictors used in the model alongside the goodness of fit. To determine what the adjusted R^2^ would be at chance, an average of adjusted R^2^ values was taken from a regression run a thousand times with randomly shuffled predictors. The Wilcoxon signed rank test was performed to see whether the adjusted R^2^ across sessions per epoch were significantly greater than the adjusted R^2^ at chance. The percentage of significant predictors was found for each regression model by dividing the number of significant predictors by the total number of predictors. The Kruskal-Wallis test with Dunn-Sidak post-hoc comparisons was done to compare the number of predictors, adjusted R^2^, and percentage of significant predictors across sessions between the onset, sustained, and offset epochs.

## References

1. Witney, A. G., Wing, A., Thonnard, J. L. & Smith, A. M. The cutaneous contribution to adaptive precision grip. Trends in Neurosciences vol. 27 637–643 Preprint at 10.1016/j.tins.2004.08.006 (2004).

2. Monzée, J., Lamarre, Y. & Smith, A. M. The effects of digital anesthesia on force control using a precision grip. J Neurophysiol 89, 672–683 (2003).

3. Augurelle, A. S., Smith, A. M., Lejeune, T. & Thonnard, J. L. Importance of cutaneous feedback in maintaining a secure grip during manipulation of hand-held objects. J Neurophysiol 89, 665–671 (2003).

4. Hertenstein, M. J., Holmes, R., McCullough, M. & Keltner, D. The Communication of Emotion via Touch. Emotion 9, 566–573 (2009).

5. Hertenstein, M. J., Keltner, D., App, B., Bulleit, B. A. & Jaskolka, A. R. Touch communicates distinct emotions. Emotion 6, 528–533 (2006).

6. Spinal Cord Injury Statistical Center, N. Traumatic Spinal Cord Injury Facts and Figures at a Glance 2024 SCI Data Sheet. (2024).

7. Anderson, K. D. Targeting Recovery: Priorities of the Spinal Cord-Injured Population. J Neurotrauma 21, (2004).

8. Greenspon, C. M. et al. Evoking stable and precise tactile sensations via multi-electrode intracortical microstimulation of the somatosensory cortex. Nat Biomed Eng (2024) doi:10.1038/s41551-024-01299-z.

9. Fifer, M. S. et al. Intracortical Microstimulation Elicits Human Fingertip Sensations. Preprint at 10.1101/2020.05.29.20117374 (2020).

10. Flesher, S. N. et al. Intracortical Microstimulation of Human Somatosensory Cortex. https://www.science.org.

11. Herring, E. Z. et al. Reconnecting the Hand and Arm to the Brain: Efficacy of Neural Interfaces for Sensorimotor Restoration after Tetraplegia. Preprint at 10.1101/2023.04.24.23288977 (2023).

12. Hughes, C. L. et al. Neural stimulation and recording performance in human sensorimotor cortex over 1500 days. J Neural Eng 18, (2021).

13. Bjånes, D. A. et al. Quantifying physical degradation alongside recording and stimulation performance of 980 intracortical microelectrodes chronically implanted in three humans for 956-2246 days. medRxiv (2024) doi:10.1101/2024.09.09.24313281.

14. Christie, B. P. et al. ‘Long-term stability of stimulating spiral nerve cuff electrodes on human peripheral nerves’. J Neuroeng Rehabil 14, (2017).

15. Tan, D. W., Schiefer, M. A., Keith, M. W., Anderson, J. R. & Tyler, D. J. Stability and selectivity of a chronic, multi-contact cuff electrode for sensory stimulation in human amputees. J Neural Eng 12, (2015).

16. Tan, D. W. et al. A neural interface provides long-term stable natural touch perception. Sci Transl Med 6, (2014).

17. Charkhkar, H. et al. High-density peripheral nerve cuffs restore natural sensation to individuals with lower-limb amputations. J Neural Eng 15, (2018).

18. Bensmaia, S. J., Tyler, D. J. & Micera, S. Restoration of sensory information via bionic hands. Nature Biomedical Engineering vol. 7 443–455 Preprint at 10.1038/s41551-020-00630-8 (2023).

19. Graczyk, E. L. et al. The Neural Basis of Perceived Intensity in Natural and Artificial Touch. http://stm.sciencemag.org/.

20. Schiefer, M., Tan, D., Sidek, S. M. & Tyler, D. J. Sensory feedback by peripheral nerve stimulation improves task performance in individuals with upper limb loss using a myoelectric prosthesis. J Neural Eng 13, (2015).

21. Kim, K. A review of haptic feedback through peripheral nerve stimulation for upper extremity prosthetics. Current Opinion in Biomedical Engineering vol. 21 Preprint at 10.1016/j.cobme.2022.100368 (2022).

22. Saal, H. P. & Bensmaia, S. J. Biomimetic approaches to bionic touch through a peripheral nerve interface. Neuropsychologia 79, 344–353 (2015).

23. Hamid, S. & Hayek, R. Role of electrical stimulation for rehabilitation and regeneration after spinal cord injury: An overview. European Spine Journal 17, 1256–1269 (2008).

24. Niloy, B., Peckham, ; & Hunter, P. Peripheral Nerve Stimulation for Restoration of Motor Function. Journal of Clinical NeurophysiologyJournal of Clinical Neurophysiology vol. 14 (1997).

25. Peckham, P. H. et al. Efficacy of an implanted neuroprosthesis for restoring hand grasp in tetraplegia: A multicenter study. Arch Phys Med Rehabil 82, 1380–1388 (2001).

26. Graczyk, E. L., Christie, B. P., He, Q., Tyler, D. J. & Bensmaia, S. J. Frequency Shapes the Quality of Tactile Percepts Evoked through Electrical Stimulation of the Nerves. Journal of Neuroscience 42, 2052–2064 (2022).

27. Valle, G. et al. Comparison of linear frequency and amplitude modulation for intraneural sensory feedback in bidirectional hand prostheses. Sci Rep 8, (2018).

28. Petrini, F. M. et al. Six-Month Assessment of a Hand Prosthesis with Intraneural Tactile Feedback. Ann Neurol 85, 137–154 (2019).

29. Delhaye, B. P., Long, K. H. & Bensmaia, S. J. Neural Basis of Touch and Proprioception in Primate Cortex. Compr Physiol 8, 1575–1602 (2018).

30. Johansson, R. S. & Vallbo, A. B. Tactile Sensory Coding in the Glabrous Skin of the Human Hand. (1983).

31. Formento, E., D’Anna, E., Gribi, S., Lacour, S. P. & Micera, S. A biomimetic electrical stimulation strategy to induce asynchronous stochastic neural activity. J Neural Eng 17, (2020).

32. Pei, Y. C., Denchev, P. V., Hsiao, S. S., Craig, J. C. & Bensmaia, S. J. Convergence of submodality-specific input onto neurons in primary somatosensory cortex. J Neurophysiol 102, 1843–1853 (2009).

33. Tanner, J., Keefer, E., Cheng, J. & Helms Tillery, S. Dynamic peripheral nerve stimulation can produce cortical activation similar to punctate mechanical stimuli. Front Hum Neurosci 17, (2023).

34. Raspopovic, S. et al. Bioengineering: Restoring natural sensory feedback in real-time bidirectional hand prostheses. Sci Transl Med 6, (2014).

35. Callier, T., Suresh, A. K. & Bensmaia, S. J. Neural Coding of Contact Events in Somatosensory Cortex. Cerebral Cortex 29, 4613–4627 (2019).

36. Saxena, S. & Cunningham, J. P. Towards the neural population doctrine. Current Opinion in Neurobiology volj. 55 103–111 Preprint at 10.1016/j.conb.2019.02.002 (2019).

37. Hayes, K. C. et al. Clinical and electrophysiologic correlates of quantitative sensory testing in patients with incomplete spinal cord injury. Arch Phys Med Rehabil 83, 1612–1619 (2002).

38. Callier, T., Gitchell, T., Harvey, M. A. & Bensmaia, S. J. Disentangling temporal and rate codes in primate somatosensory cortex. The Journal of Neuroscience e0036242024 (2024) doi:10.1523/jneurosci.0036-24.2024.

39. Suresh, A. K. et al. Sensory computations in the cuneate nucleus of macaques. 118, (2021).

40. Gorman, P. H. & Mortimer, J. T. The Effect of Stimulus Parameters on the Recruitment Characteristics of Direct Nerve Stimulation. IEEE TRANSACTIONS ON BIOMEDICAL ENGINEERING vol. 30 (1983).

41. Principles of Neuroscience.

42. McCreery, D., Cogan, S., Kane, S. & Pikov, V. Correlations between histology and neuronal activity recorded by microelectrodes implanted chronically in the cerebral cortex. J Neural Eng 13, (2016).

43. Hutchison, B. et al. Perceptual Differences between Cortical and Peripheral Stimulation Strategies for Sensory Restoration. Preprint at 10.1101/2025.08.20.25334094 (2025).

44. George, J. A., et al. Biomimetic Sensory Feedback through Peripheral Nerve Stimulation Improves Dexterous Use of a Bionic Hand. http://robotics.sciencemag.org/ (2019).

45. Valle, G. et al. Biomimetic Intraneural Sensory Feedback Enhances Sensation Naturalness, Tactile Sensitivity, and Manual Dexterity in a Bidirectional Prosthesis. Neuron 100, 37–45.e7 (2018).

46. Delion, A. L. The Moving Two-Point Discrimination Test: Clinical Evaluation of the Quickly Adapting Fiber/Receptor System. (1978).

47. Semmes, J., Weinstein, S., Ghent, L. & Teuber, H.-L. Somatosensory Changes after Penetrating Brain Wounds in Man. Somatosensory changes after penetrating brain wounds in man. (Harvard Univer. Press, Oxford, England, 1960).

48. Shannon, R. V. A Model of Safe Levels for Electrical Stimulation. IEEE Trans Biomed Eng 39, 424–426 (1992).

49. Young, D. et al. Signal processing methods for reducing artifacts in microelectrode brain recordings caused by functional electrical stimulation. J Neural Eng 14, 535–562 (2018).

