## Supplementary Information for "Cortical representation of sensation elicited by peripheral nerve stimulation in an individual with incomplete spinal cord injury"

### **Supplementary Materials**

|  | Normal | Thumb | Index | Middle | Ring | Little | Thenar Eminence | Back of the Hand |
| --- | --- | --- | --- | --- | --- | --- | --- | --- |
| Semmes-Weinstein Monofilament Test | 2.44- 2.83 | 4.31 | 4.17 | 4.17 | 4.17 | 4.17 | 4.56 | 6.65 |
| Static Two-Point Discrimination Test (mm) | 2-5 | 12 | 7 | 7 | 5 | 5 | N/A | N/A |

Supplemental Table 1. Sensory assessments completed after the implantation of the peripheral nerve electrodes and cortical arrays. Results from both the Semmes-Weinstein Monofilament Test and Static Two-Point Discrimination Test demonstrate that the participant has residual sensation on all fingers and that the thumb has poorer sensory function than the other fingers.

| **Days Post Implant** | **Contact** | **Trials Collected** | **Low FR Trials** | **High FR Trials** | **Highly Correlated Trials** | **% Trials Retained** | **% Elec Retained** |
| --- | --- | --- | --- | --- | --- | --- | --- |
| 308 | M2 | 149 | 0 | 0 | 0 | 100% | 66% |
| 318 | M2 | 145 | 38 | 7 | 22 | 59% | 100% |
| 400 | M1 | 148 | 0 | 0 | 0 | 100% | 55% |
| 434 | M1 | 116 | 0 | 0 | 1 | 99% | 100% |
| 462 | M1 | 88 | 0 | 0 | 0 | 100% | 48% |
| 562 | U5 | 100 | 41 | 0 | 19 | 40% | 100% |
| 669 | U5 | 159 | 47 | 0 | 55 | 36% | 100% |
| 674 | U5 | 155 | 6 | 1 | 33 | 74% | 100% |
| 795 | U5 | 150 | 9 | 0 | 10 | 87% | 95% |
| 801 | U4 | 150 | N/A | N/A | N/A | N/A | N/A |
| 827 | U4 | 240 | 27 | 0 | 1 | 88% | 100% |
| 828 | U4 | 220 | 21 | 0 | 2 | 90% | 100% |
| 1122 | U5 | 179 | N/A | N/A | N/A | N/A | N/A |
| 1128 | U5 | 150 | 0 | 0 | 0 | 100% | 52% |

Supplemental Table 2. Dataset used in the study. Rows in white are sessions where only perceptual data was collected due to improper setup of the recording equipment. Rows in grey are sessions that were excluded due to a low trial retention of under 50% caused by poor neural recordings. Trials were excluded if they had too low or high of a firing rate, or if the neural activity in the trial was highly correlated across electrodes (see Methods for details). Note that all the trials with too high of a firing rate were also highly correlated. Rows with both perceptual and neural data are color coded based on their corresponding PNS input (teal = M1, blue = M2, orange = U4, pink = U5).

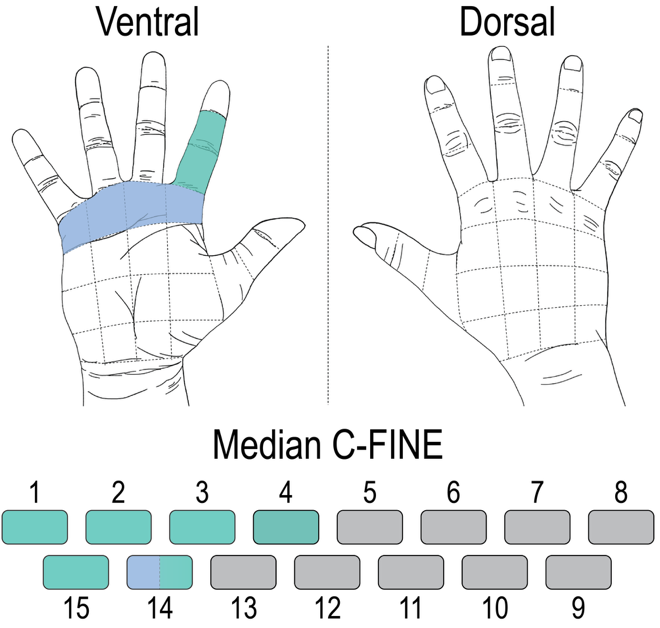

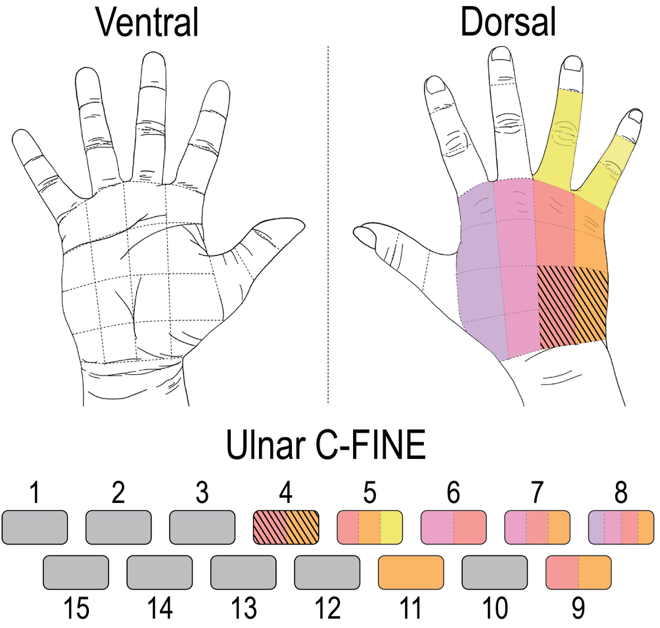

Supplemental Figure 1. Projected fields resulting from delivering PNS at sensory threshold on C-FINE contacts on the median nerve (left) and ulnar nerve (right). Electrode mapping surveys on the median and ulnar nerve were conducted 343 days and 525 days post-implant, respectively. The regions of the hand at which each contact elicited sensation are color-coded, and grey indicates that stimulating the contact did not produce a sensory response (though it may have elicited a motor response).
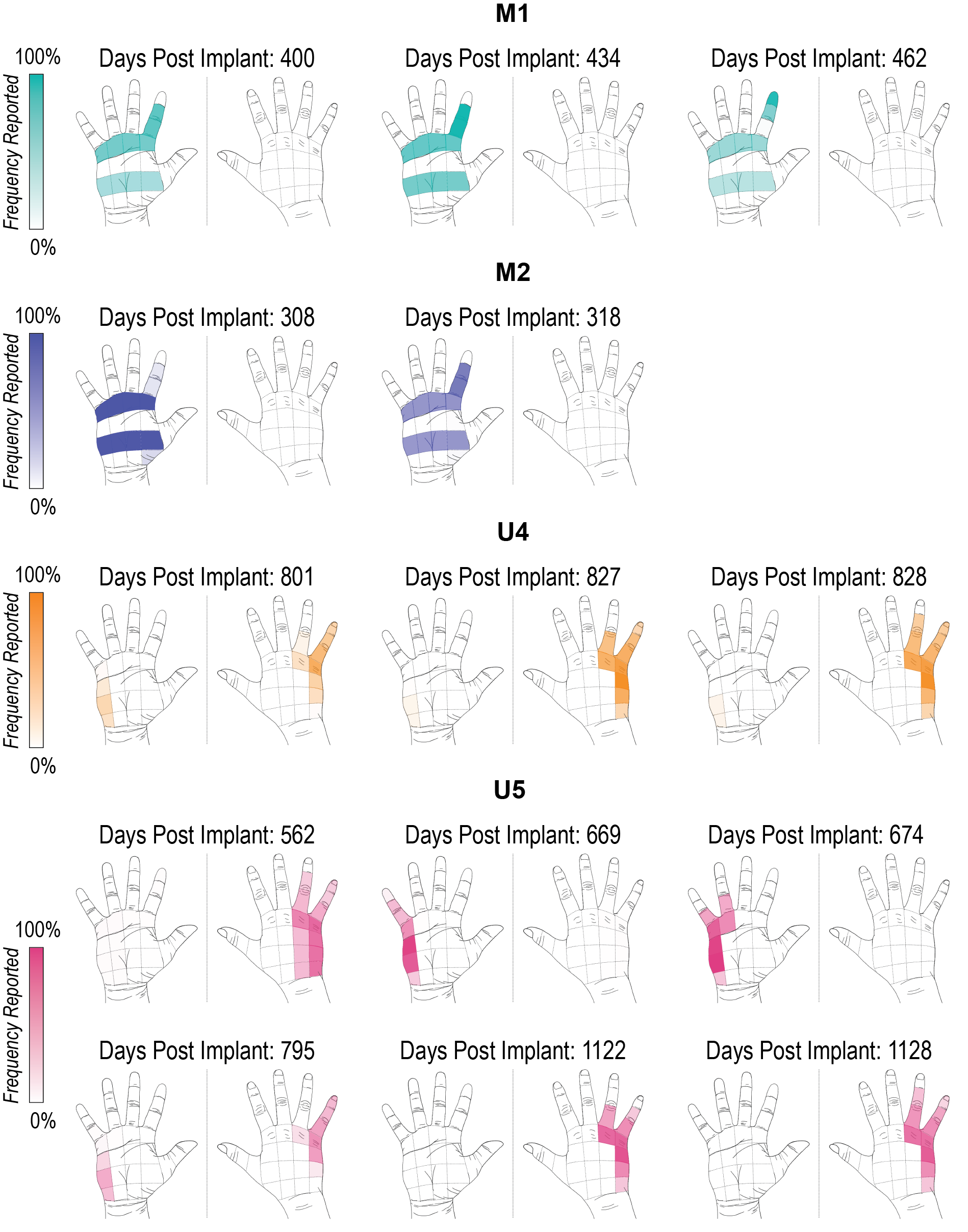
Supplemental Figure 2. Reliability of the projected fields generated from the four PNS contacts selected for this study. M1 and M2 are two contacts on the median nerve, while U4 and U5 are two contacts on the ulnar nerve. Projected fields are shown for each session, and color corresponds to the PNS contact. Opacity of each hand region indicates the frequency at which it was reported across perceivable trials per session (n = 43 to 150 perceivable trials, depending on the session).

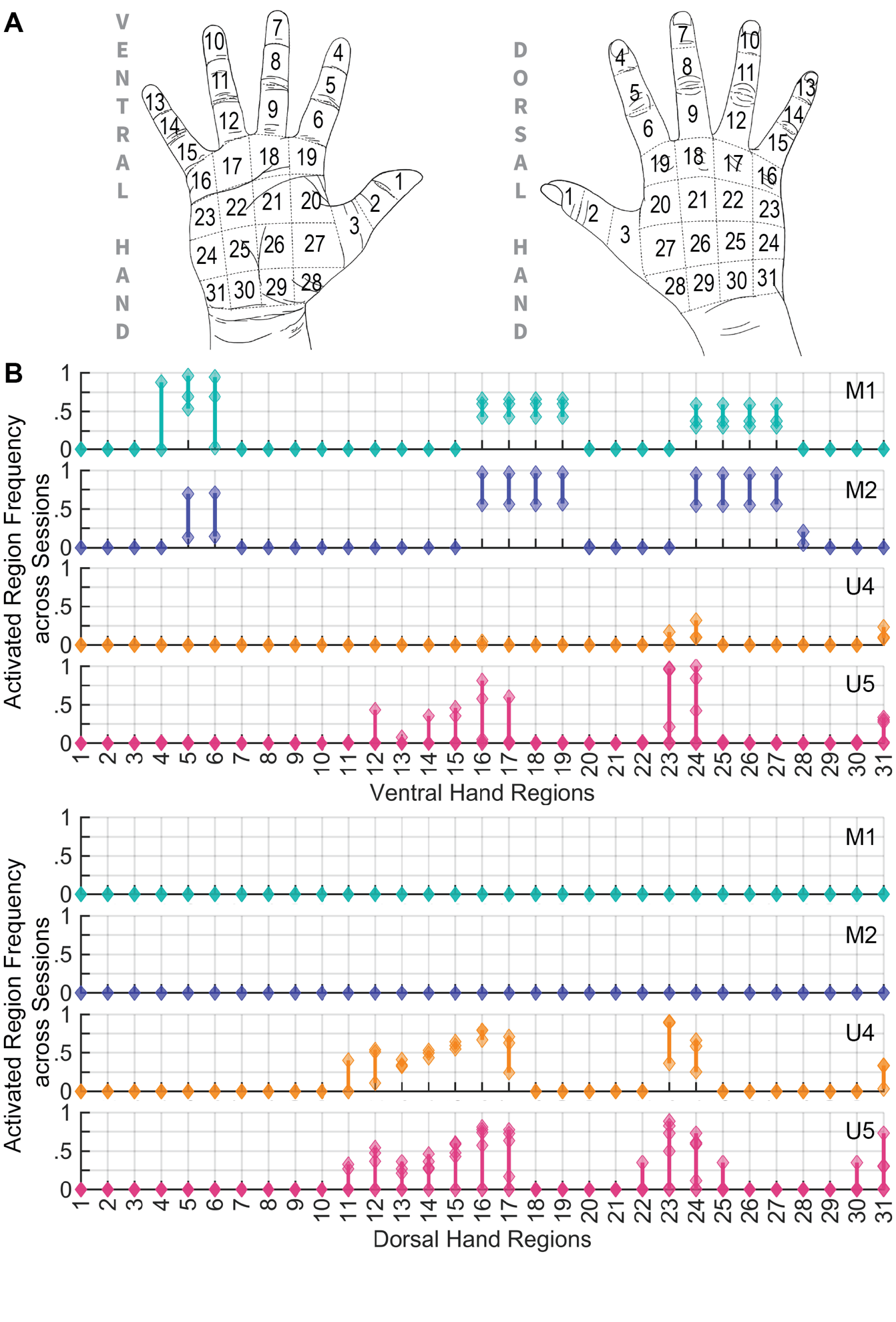

Supplemental Figure 3. A) The hand diagram displayed for the participant to reference when reporting projected field locations during PNS. The participant verbally reported the discrete regions on the hand via their numerical labels. Verbal reports were necessary because the participant has complete paralysis of all four limbs and thus could not draw on a hand diagram. B) The consistency of how often a hand region was felt across sessions for a given PNS contact. For a hand region on either the dorsal or ventral side, each diamond represents how often it was reported during a session. The vertical line indicates the range between the frequencies of the region being reported across sessions (n = 3 sessions for M1, n = 2 sessions for M2, n = 3 sessions for U4, and n = 6 sessions for U5). Hand regions with a narrow range indicate that the frequency of the region being reported was consistent across sessions, whereas regions with a wider range indicate higher variability of the region being reported. Note that several regions for M1 had a wide range because completely different parts of the index finger were felt in different sessions. Most regions for U5 had a wide range because the projected fields alternated between the dorsal and ventral sides of the hand across sessions.

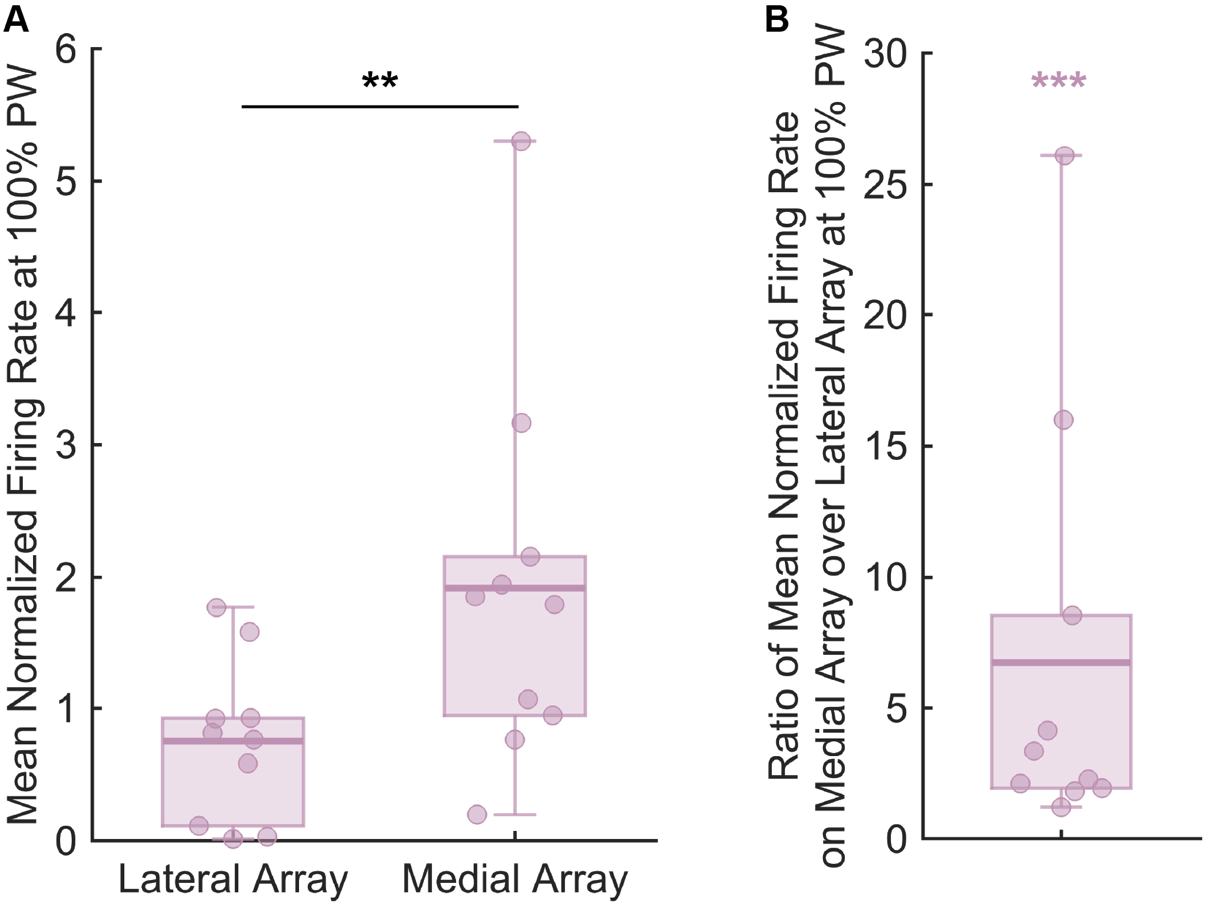

Supplemental Figure 4. Comparison of cortical activation recorded on the lateral and medial arrays in primary somatosensory cortex. A) Mean normalized firing rate during the onset epoch for the 100% PW condition across all sessions (n = 10 sessions). There was significantly higher activation on the medial array than the lateral array. Black ‘**’ indicates a significant difference of p < 0.01 using the rank-sum test. B) Ratio of the mean normalized firing rate at 100% PW during onset on the medial array over the mean normalized firing rate at 100% PW during onset on the lateral array across all sessions (n = 10 sessions). Purple ‘*’ indicates that the fractions are significantly greater than 1 (right-sided Wilcoxon signed rank test, p < 0.001).

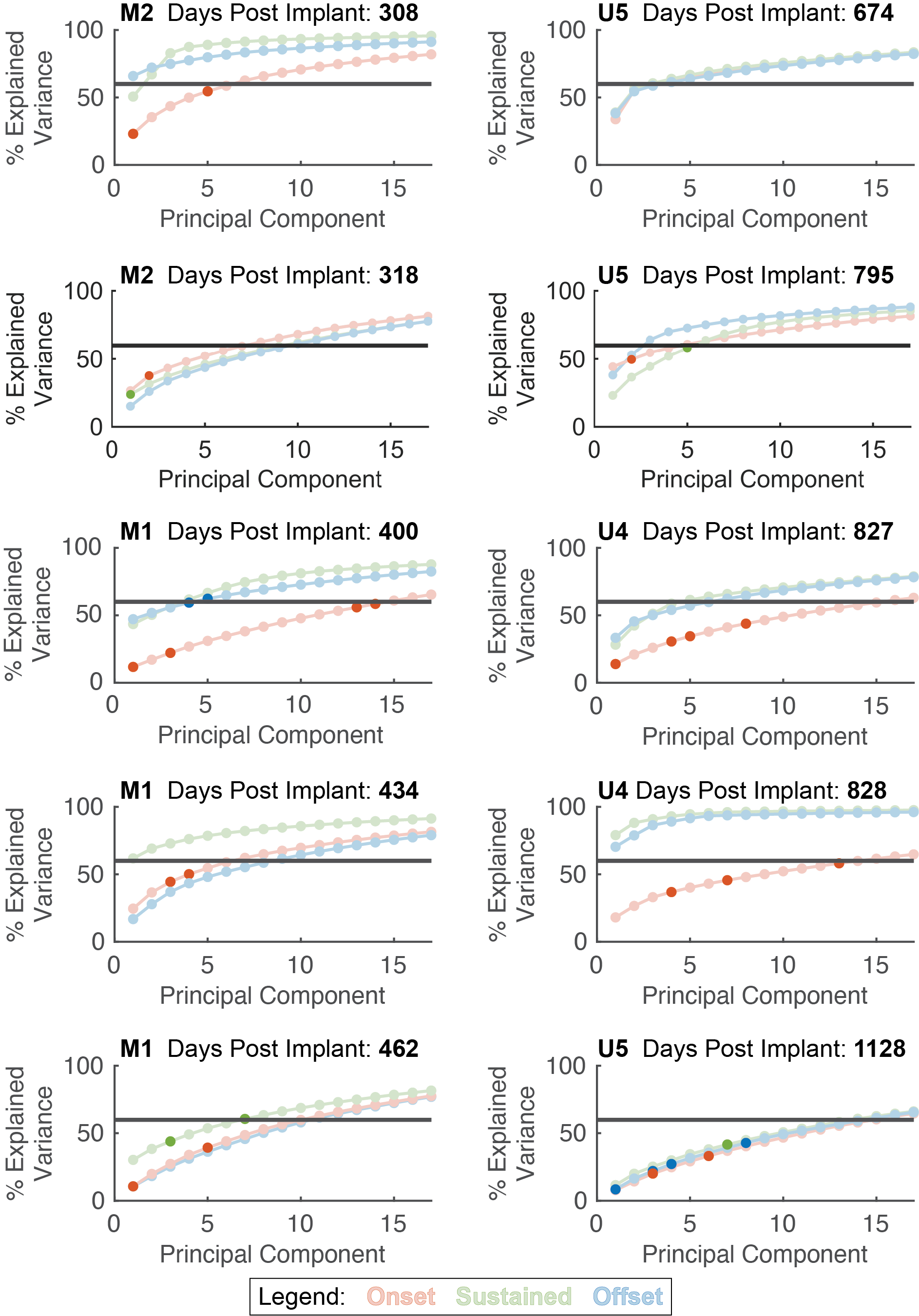

Supplemental Figure 5. Cumulative percentage of variance explained from principal components of normalized firing rates averaged per epoch per session. The red, green, and blue colors represent the onset, sustained, and offset epochs respectively, and the circles denote the cumulative percentage of variance for the given principal component. For each epoch per session, a multiple regression model predicting perceived intensity was constructed from the principal components that cumulatively accounted for 60% of the variance (grey horizontal line). Circles filled with a darker color denote the principal components that were significant in the regression between that epoch and perceived intensity for that sessions’ model. Note that onset (red) had more significant predictors than the models for the other epochs.
